# Respiratory support with Continuous Positive Airway Pressure in preterm neonates: an analysis of coverage and quality of care in 66 neonatal units in Kenya, Malawi, Nigeria and Tanzania implementing with the NEST360 Alliance

**DOI:** 10.64898/2026.06.20.26356142

**Authors:** Kristina Shemwell, John Wainaina, Joy E. Lawn, Nahya Salim Masoud, Rebecca E. Penzias, Lucas Malla, Mariam Johari, Robert Tillya, Christine A. Bohne, Msandeni Chiume, Samuel K. Ngwala, Olabisi O. Dosunmu, Chinyere Ezeaka, George Okello, William M. Macharia, Natasha R. Rhoda, Edith Gicheha, Nebiyou Hailemariam, Morris Ondieki Ogero, Junwei Chen, Eric O. Ohuma, Rebecca Richards-Kortum, Maria Oden, James H. Cross, Kondwani Kawaza, Elizabeth M. Molyneux, With the NEST360 Neonatal Inpatient Dataset and Data Systems Collaborative Group, Context Tracker and the NEST360 Health Facility Assessment Collaborative Group

**Author notes:** Corresponding Author: Kristina Shemwell. Co-first authors (Kristina Shemwell, John Wainaina). Co-senior authors (James H. Cross, Kondwani Kawaza, Elizabeth M. Molyneux).

## Abstract

**Background:** Prematurity is the leading cause of child deaths worldwide, with the highest neonatal mortality in sub-Saharan Africa. Respiratory distress syndrome (RDS) is the leading mortality pathway in preterm neonates, but continuous positive airway pressure (CPAP) has high impact. This analysis reports CPAP coverage and quality-of-care for preterm neonates admitted to 66 neonatal units in Kenya, Malawi, Nigeria and Tanzania.

**Methods:** Analyses used individually-linked neonatal inpatient data and cross-sectional health systems data. All admitted neonates were eligible for inclusion (January 2021–December 2024). Service readiness for CPAP delivery and mean CPAP coverage were described for CPAP-eligible newborns (weighing <1500G and symptomatic newborns >1500g). Quality-of-care cascades were constructed to illustrate key indicators. Survival among CPAP-eligible neonates was analysed using regression models, stratified by clinical severity scores.

**Results:** 375,255 newborn admissions were analysed in 66 neonatal units. Functional CPAP availability varied with median 16% of days (IQR: 4-47%) classified as high demand (>1.5 eligible newborns per CPAP). Of 64,761 CPAP-eligible neonates, 22,006 (34%, 95% CI 33-34%) received CPAP. All countries showed improvement in CPAP coverage, with Tanzania’s hospitals recording 63% increase in mean coverage (p-value=0.001) over time. Quality-of-care cascades showed treatment was initiated <24 hours after birth and continued for >1 day for 42% (95% CI 41-43%) of eligible neonates receiving CPAP. Only 10% of neonates <1500g started CPAP within the first hour of life. Among newborns on CPAP, 55% also received KMC (from 48% in Tanzania to 88% in Nigeria). Among newborns with high clinical severity, those treated with CPAP had a higher probability of survival (32%, 95% CI 29–36%) than those who were not (23%, 95% CI 21–26%). Odds of survival were higher for CPAP-eligible newborns whose mothers received antenatal corticosteroids (aOR 1.07, p=0.001). Lower aOR of survival was associated with hypoglycaemia (aOR 0.71, p<0.001), respiratory distress (aOR 0.91, p<0.001), and outborn newborns (aOR 0.72, p<0.001).

**Conclusion:** CPAP coverage and quality are critical for premature neonates. Clinical cascades highlight quality gaps, particularly in timely prophylactic CPAP initiation and appropriate duration. Improving comprehensive care quality for newborns on CPAP, including provision of co-interventions and maternal antenatal corticosteroids, can improve survival for preterm neonates.

**KEY FINDINGS:** *WHAT WAS KNOWN?:*

- Respiratory distress syndrome (RDS) is the leading pathway to mortality in preterm neonates, but continuous positive airway pressure (CPAP) is a high-impact intervention and recommended by WHO.
- There are health systems barriers to CPAP implementation in low- and middle-income country (LMIC) settings. No current published reports analyse large-scale coverage of CPAP in neonatal units across sub–Saharan African contexts or report on quality of care.
- There is a gap in published clinical severity scores in LMICs which can be used in statistical analyses to predict survival.
- NEST360 Alliance is supporting governments to implement small and sick newborn care in hospitals in Malawi (national scale), Kenya, Tanzania, and Nigeria, with a focus on high-impact interventions such as CPAP.

*WHAT WAS DONE THAT IS NEW?:*

- This multi-site analysis described service readiness, coverage, and care quality for over 370,000 inpatient neonates in 66 neonatal units, using individually-linked clinical data routinely collected from newborn admission records.
- Individual-level data were used to assess CPAP quality-of-care with novel quality-of-care cascades, enabling visualisation of quality at multiple steps in the care pathway during neonatal admissions.
- A clinical severity score predicting survival was developed to understand outcomes among CPAP-eligible neonates.

*WHAT WAS FOUND?:*

- Limited CPAP equipment availability was demonstrated by nearly one-third (31%) of days having ≥1.5 eligible babies per device.
- CPAP coverage increased over time in all countries, with the largest increase in Tanzania from 25% to 88% over three years.
- CPAP quality-of-care cascades pooled across all sites showed 42% of eligible neonates who received CPAP were initiated within the first day of life and continued for at least one day. Among neonates under 1500g, only 10% started CPAP within the first hour of life.
- High clinical severity leads to death without CPAP (predicted probability of survival 23%, 95% CI 21–26%), but survival improves with CPAP (predicted probability of survival 32%, 95% CI 29–36%).

*WHAT NEXT?:*

- Both CPAP coverage and quality are critical for premature neonates. Neonatal units in similar high-burden settings can apply clinical care cascades to improve care quality for the right babies at the right time.
- The respiratory care package also includes antenatal corticosteroids and prevention/ management of apnoea of prematurity. Stronger linkages between maternal and newborn care would improve this package. Future analyses could assess co-coverage with this more comprehensive respiratory care bundle.

## BACKGROUND

Globally, around half (47%) of deaths among children under five years occur during the neonatal period (day 0-28) [1–3], resulting in an estimated 2.3 million neonatal deaths in 2023 [4]. The leading cause of child deaths is direct complications from preterm birth [1–3]. Neonatal mortality is inequitably distributed, with an estimated 79% of deaths concentrated in sub-Saharan Africa and South Asia [2, 5], where there is also a high prevalence of preterm births [6]. Despite reduction in under-five mortality rates globally, reduction in neonatal mortality has been slower, particularly in these regions [5]. Sub-Saharan Africa has the highest burden with a neonatal mortality rate (NMR) of 26 deaths per 1,000 live births in 2023 – far from the SDG3.2 target of 12 or fewer neonatal deaths per 1000 live births [2]. Newborns have a varying range of clinical severities which can predict their risk of mortality, which can be addressed using clinical severity scores. Several clinical severity scores for predicting mortality or illness severity are published, but few across the weight spectrum for LMICs which can be used for statistical analyses, such as the NMR-2000 score [7–12].

Respiratory distress syndrome (RDS) is the leading cause of mortality for preterm neonates [13, 14], and high prevalence rates and case fatality rates (estimated 52.5% fatality) have been reported in sub-Saharan Africa [15, 16]. Hence to accelerate progress the prevention and care of RDS must be improved [17].

Continuous positive airway pressure (CPAP), is an intervention recommended by WHO [18, 19] for management of RDS. CPAP’s positive effects on survival have been demonstrated in low-and-middle-income country (LMIC) settings [20, 21]. Studies of CPAP use from Nigeria, Tanzania, Kenya, and Malawi all demonstrated significant improvements in survival of premature neonates after implementation of CPAP [22–27]. A systematic review of observational studies also found a 66% reduction of in-hospital mortality among preterm infants receiving CPAP [28].

A recent Cochrane review analysing randomized controlled trials of prophylactic CPAP found improved survival, less need to escalate to higher level-of-care such as mechanical ventilation, and fewer pulmonary complications in preterm infants who received prophylactic CPAP compared to more invasive respiratory support [29, 30]. Due to gaps in data for gestational age in many LMIC settings, birthweight is often used as a surrogate marker for prematurity to guide eligibility for CPAP initiation [24].

Despite evidence for CPAP, there are barriers to CPAP coverage and quality especially in high-burden settings – for example lack of equipment, varying types of CPAP devices and frequent use of improvised devices [31–33]. Human resources for health were also identified as a major constraint in another study assessing health systems bottlenecks [34]. There are limited data on coverage of CPAP in neonatal units in high-burden countries, though a recent survey of clinicians opinions across 49 countries in Africa suggested that CPAP use may be highest in larger cities, private facilities, and the most well-equipped government hospitals, where up to 67% of such facilities were reported to have provided CPAP [35]. However, we found no current published reports with analyses of large-scale coverage or quality of CPAP in neonatal units across sub-Saharan Africa.

### Aim and objectives

This paper is part of a supplement reporting findings and learnings of the Newborn Essential Solutions and Technologies (NEST360) Alliance, including 23 organizations, working with five African governments (Malawi- in all government hospitals; Kenya, Tanzania, Nigeria, and more recently Ethiopia), to reduce neonatal inpatient deaths by improving small and sick newborn care (SSNC) in hospitals through device installation, education, and data-driven quality improvement. We aimed to describe CPAP service readiness, coverage, quality-of-care, and outcomes for neonates across 66 neonatal units in four African countries implementing with NEST360: Malawi, Kenya, Tanzania, and Nigeria.

### Objectives

**1. Service readiness**: Describe hospital-level factors which may impact care of preterm neonates with CPAP, including neonatal unit capacity, device and staffing ratios, and other health systems factors, using hospital-level data.
**2. CPAP coverage**: Describe CPAP coverage over time at country population-level for admitted neonates eligible to receive CPAP in hospitals analysed, using aggregate data.
**3. CPAP quality-of-care**: Illustrate quality of CPAP care through cascades including key quality indicators using individual-level data.
**4. Outcomes for CPAP**: Analyse survival among CPAP-eligible neonates according to clinical severity using individual-level data.

## METHODS

### Overview

A multi-site cross-sectional observational design using secondary data analysis was used. Data were collected routinely at each site for all newborn admissions during the analysis period of January 2021 through December 2024. Hospital-level data were collected using cross-sectional questionnaires in 2023 and a weekly hospital-level context tracker from 2024. This analysis was reported following the Strengthening the Reporting of Observational Studies in Epidemiology (STROBE) checklist (see **Additional File 1**) [36].

### Study Setting

This analysis focuses on hospitals in four of the five countries (Malawi, Kenya, Tanzania, and Nigeria) working with the NEST360 alliance. It includes neonatal units which completed initial device and dataset implementation and have ongoing quality improvement and training activities (excluding data prior to implementation). Prospective data collection for all newborn admissions is ongoing for implementing sites [37]. Inpatient data were available in 64 hospitals, two of which have geographically separated inborn and outborn units, accounting to 66 neonatal units. One hospital in Malawi completed initial facility assessment but did not proceed with implementation (65 hospitals assessed for service readiness).

### Data Sources

The Newborn Inpatient Dataset (NID) is a co-designed core inpatient neonatal dataset which aims to improve care quality using parsimonious data on every neonatal admission including clinical risk characteristics, inpatient care and outcomes [37]. Trained local data clerks enter data from medical records into REDCap, a web-based application.

The NID in Malawi is the government data source; and in Tanzania and Nigeria, the NID is in process of government linkage. In Kenya, variables are exported from NID to a research project data system, the Clinical Information Network (CIN) dataset [38], which has less focus on QI so has some missing variables on coverage and quality of care compared to NID.

The NEST360/UNICEF Health Facility Assessment (HFA) for small and sick newborn care provides a standardised interviewer-led assessment of facility resources with observation to assess service readiness in each hospital based on health systems building blocks [39]. To assess contextual indicators (e.g., occupancy, device ratios, staffing), we leveraged the hospital-level Context Tracker, which was introduced in April 2024. Accordingly, contextual analyses were restricted to April 2024–March 2025 to align with the first full year of available data, while admissions analyses for other objectives used the broader 2021–2024 dataset. Data tools [37, 39–41] are available open access online and in **Additional File 7,8** [42].

### Methods by objective

#### Service readiness

Readiness was assessed using data from the following sources: the HFA (65 hospitals, 2023), Context Tracker (64 hospitals, weekly since 2024), and NID (64 hospitals, April 2024-March 2025). Context Tracker data was collected after the main analysis period but useful in describing the hospitals’ service availability. Hospitals were grouped by country; in Malawi, district and central hospitals were analyzed separately due to the country’s hospital classification and large volume of hospitals in Malawi.

##### Neonatal unit occupancy

Using neonatal unit admissions and discharge data and monthly facility-reported bed capacity from the Context Tracker, we calculated daily neonatal unit occupancy per facility. Capacity was defined as available cots and incubators; missing values were imputed using facility-specific medians across observed months. Daily occupancy (number of neonates ÷ unit capacity) was summarized monthly and reported as the median over 12-months.

##### CPAP device-to-eligible-baby ratio

Using NID and context tracker data, we estimated ratios of functional CPAP devices to neonates eligible for CPAP (defined under Objective 2). Missing device values were imputed from hospital-specific medians of non-zero months. Estimated CPAP need assumed up to four consecutive days of CPAP per eligible neonate. Inadequate CPAP device coverage was defined as days where estimated CPAP need exceeded the number of functional CPAP devices.

##### Nurse-to-baby ratios

Daily nurse-to-baby ratios were calculated using weekly staffing data from the Context Tracker and daily NID data. Ratios were computed separately for day and night shifts. We defined a high-burden shift as >8 babies per nurse, reflecting neonatal unit operational norms and consistent with prior workload assessments [41], and calculated the percentage of days exceeding this threshold.

##### Health System Building Block Scoring for CPAP

Service readiness was scored [40] based on inputs required to provide basic, comprehensive, or gold-standard respiratory support with CPAP as previously published [40]. Sub-scores assessed input needed for screening and CPAP management. Median scores were calculated per neonatal unit and by country. Overall scores were expressed as a percentage of the maximum possible score and weighted equally per component [43].

#### Coverage

CPAP coverage was analysed using a denominator of eligible neonates, stratified by eligibility criteria including birthweight groups and respiratory symptoms. Hospitals implementing with NEST360 use clinical criteria to initiate CPAP early for all neonates with a birthweight of 1000-1499g, and those weighing ≥1500g with symptoms, notably respiratory distress and/or hypoxia [44]. Neonates weighing <1000g should receive CPAP if appropriate devices are available [44] as well as symptomatic neonates with birthweight ≥2000g. Due to data source differences in Kenya, variables related to hypoxia were not available, but symptomatic criteria were used. In alignment with other neonatal analyses [45], neonates weighing <500 grams or >5500 grams were considered implausible and excluded.

Missingness in CPAP data was explored. Complete case analysis methodology was used to assess CPAP coverage for neonates eligible to receive the therapy. Quarterly mean coverage proportions were calculated at the neonatal unit-level for eligible neonates who received CPAP, then means of these mean proportions were taken on a country population-level for hospitals analysed. Due to fewer admissions in Nigeria, coverage over time was assessed annually rather than quarterly.

#### Quality of care

Individual-level data were aggregated by country to create quality of care cascades for neonates who received CPAP by adapting the individual-level clinical cascade framework used for HIV care [46–49]. Clinical quality indicators were selected based upon known clinical impact, such as timing of CPAP administration and duration [29, 30, 50–52], as well as co-administration of other high-impact interventions. The type of CPAP device used was assessed separately as these data were only collected later (June 2024-December 2024).

#### Outcomes

The outcome of interest was survival among CPAP-eligible newborns. Given the challenges with reverse mortality bias for newborns receiving CPAP, a parsimonious clinical severity score using newborn-level clinical data was created to predict survival using the smallest number of candidate variables for newborns weighing <2500 grams. This score was adapted from validated clinical severity scores for neonatal mortality [7–9], noting a gap in LMICs for score predicting survival [11, 12]. Based upon data availability, statistical strength of association for predicting survival, and three categorical levels based on WHO and other international classifications for ordinal score [53, 54], the final severity score includes gestational age, pulse oximetry on admission, and hypothermia on admission **(Table 1)**. This score was tested in the analysis dataset and a separate NID dataset for internal validity. Additional details on score development can be found in **Additional File 7**.

**Table 1.**
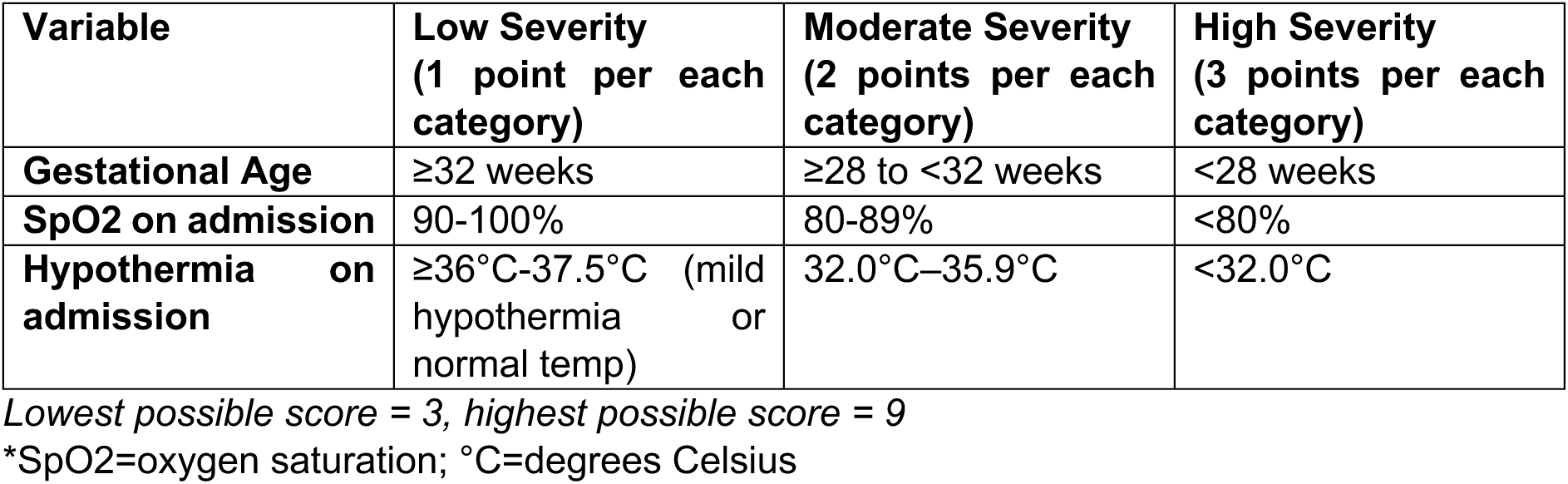
Severity score for predicting survival.

Mixed-effects regression modelling was performed to assess relationships of clinical variables with survival and account for possible confounding as well as clustering of the data by hospitals. Clinical explanatory variables were hypothesized to have impact on survival based on prior literature and expertise [23, 25, 29–32, 55–59] and had statistical association with survival in prior models **(Additional File 7)**.

## RESULTS

### Overview

Health systems data were available from Context Tracker for 64 hospitals and service readiness data from HFA for all 65 hospitals. The 65 hospitals represented in the HFA comprise 67 neonatal units, as two hospitals have separate inborn and outborn neonatal units; however, one neonatal unit did not complete implementation to allow for neonatal inpatient data. Between 1 January 2021 and 31 December 2024, 375,255 newborns were admitted across 66 neonatal units. Of these, 70,083 met criteria for CPAP eligibility **(Figure 1).** Newborn characteristics across each country were described in **Table 2**.

**Figure 1.**
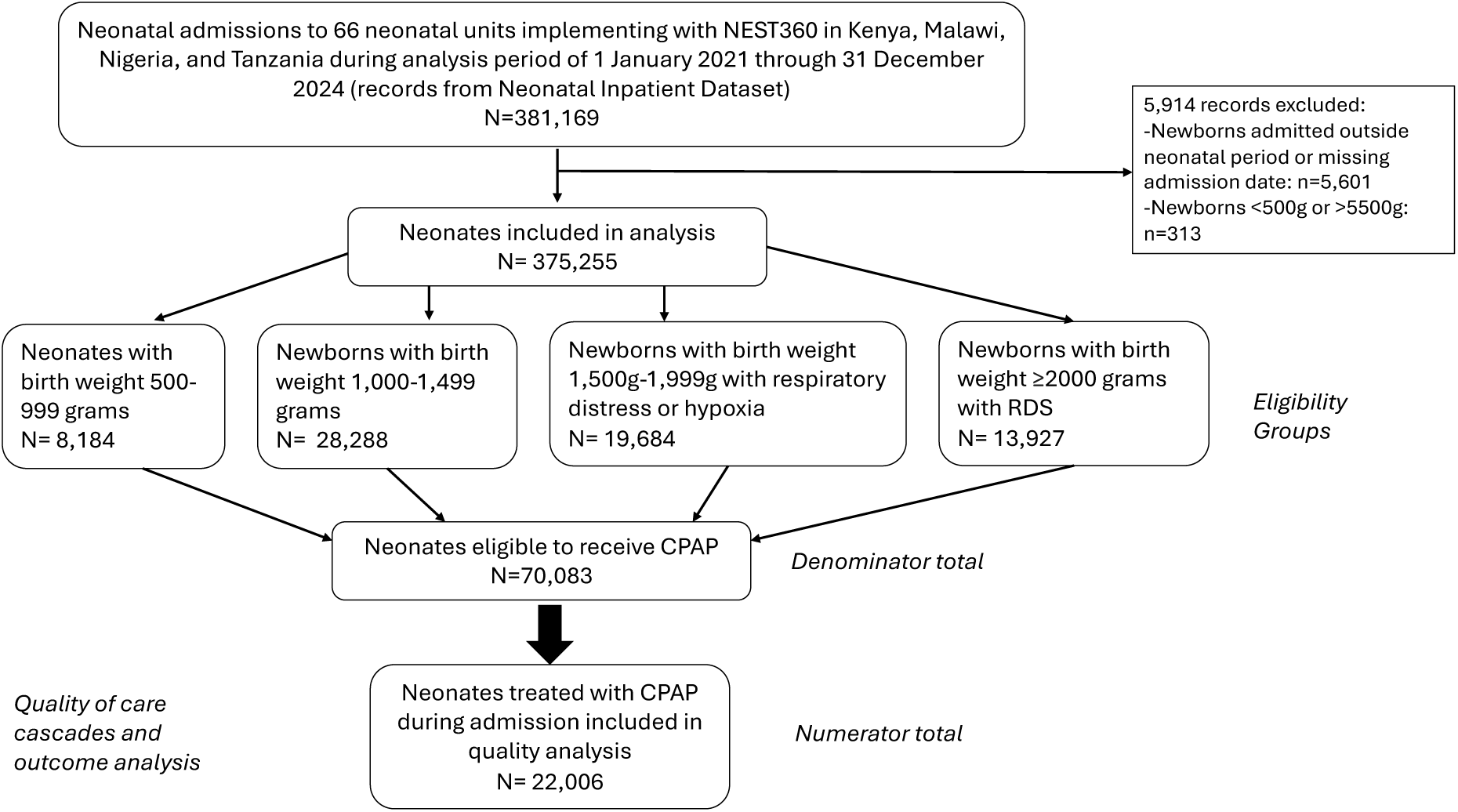
Flow diagram for the selection of newborns for this analysis (January 2021-December 2024). Analysis for CPAP coverage, quality-of-care, and outcomes at the individual-level. Only newborns admitted within the neonatal period were included (n=5,601 neonates excluded with missing admission date or admission > 28 days of life). Admitted neonates weighing <500 grams or > 5500grams were excluded (n=313 neonates).

**Table 2.**
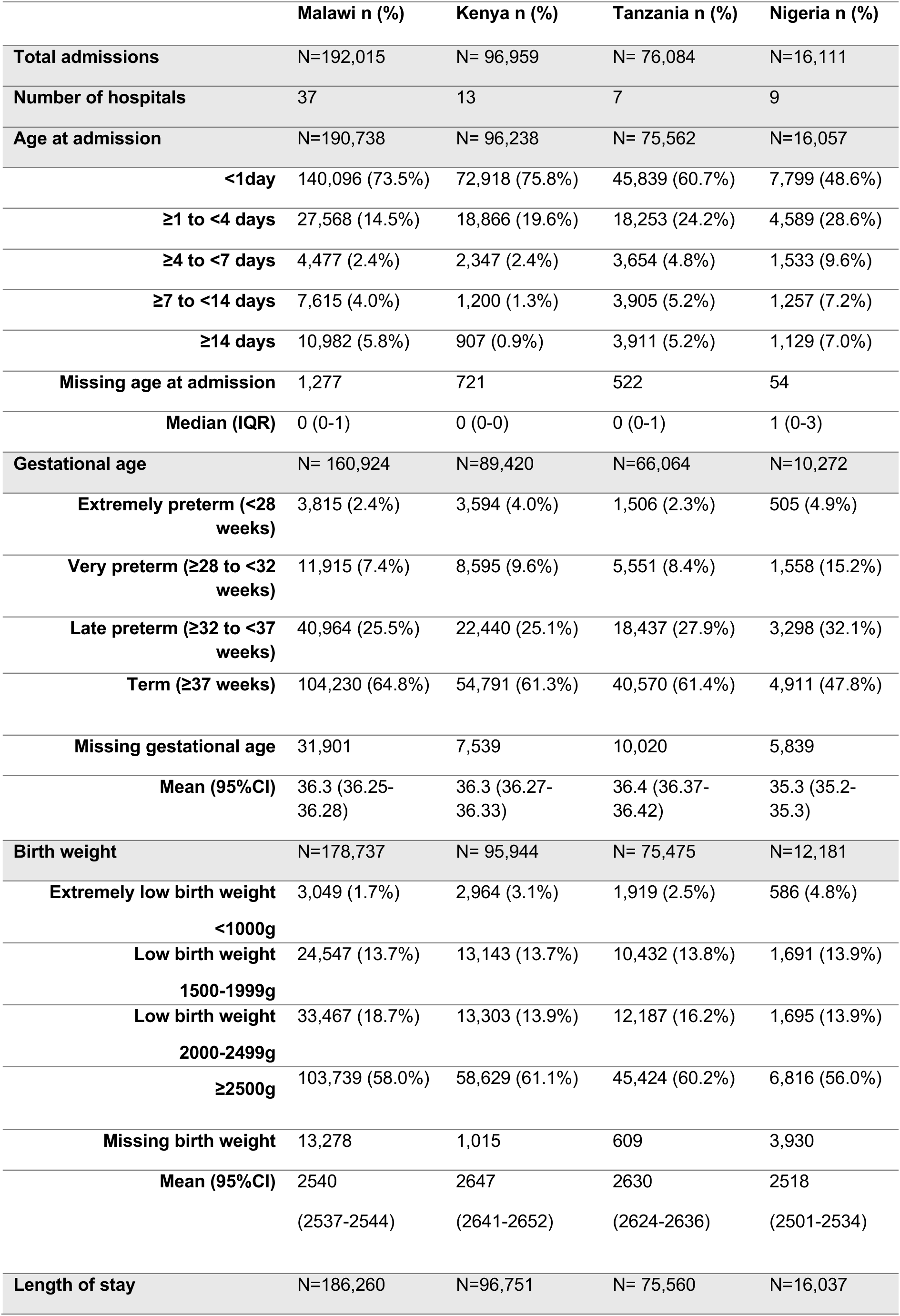
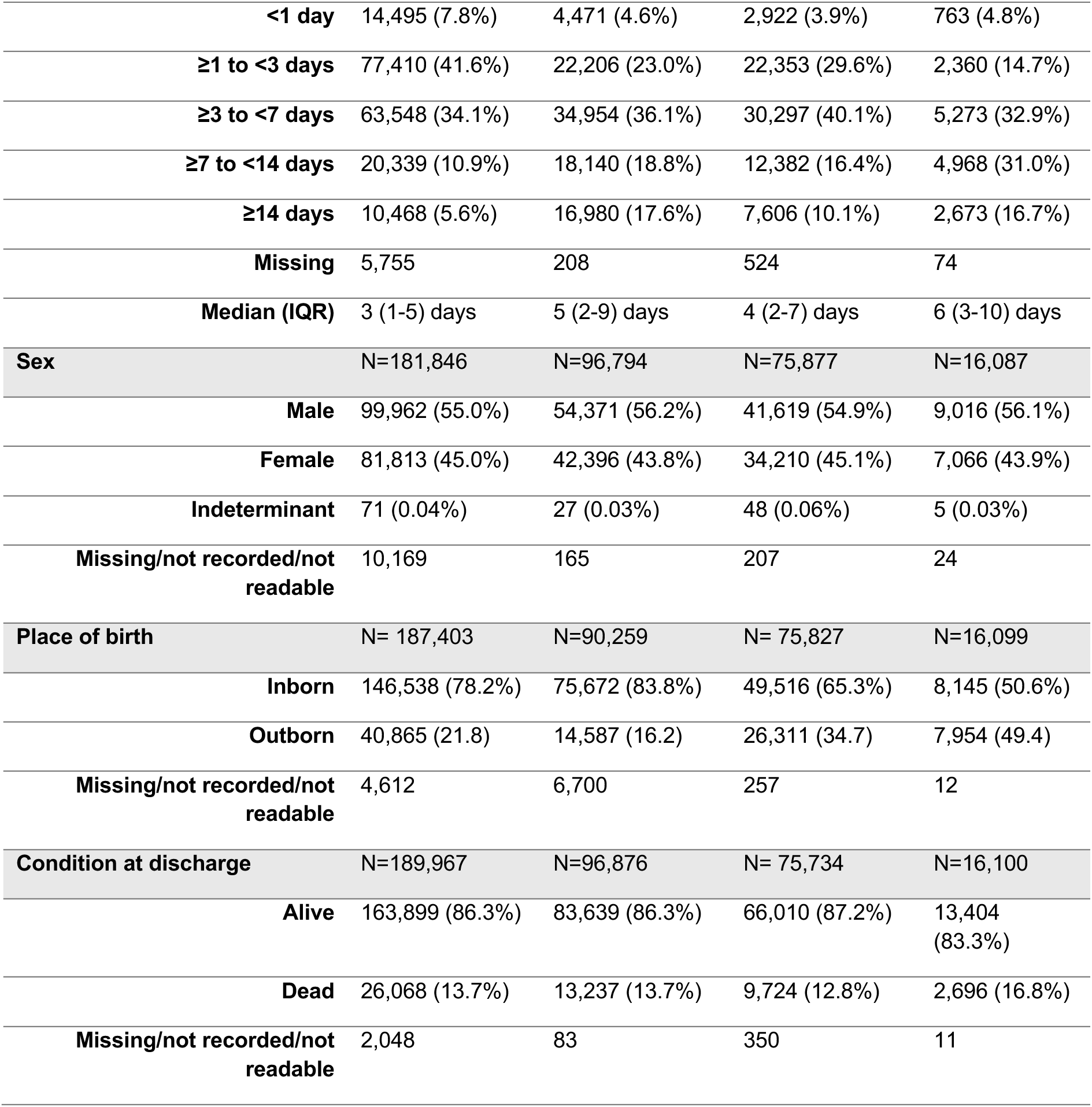
Characteristics of Admitted Neonates (January 2021-December 2024)

|  | Malawi n (%) | Kenya n (%) | Tanzania n (%) | Nigeria n (%) |
| --- | --- | --- | --- | --- |
| **Total admissions** | N=192,015 | N= 96,959 | N= 76,084 | N=16,111 |
| **Number of hospitals** | 37 | 13 | 7 | 9 |
| **Age at admission** | N=190,738 | N= 96,238 | N= 75,562 | N=16,057 |
| <1day | 140,096 (73.5%) | 72,918 (75.8%) | 45,839 (60.7%) | 7,799 (48.6%) |
| ≥1 to <4 days | 27,568 (14.5%) | 18,866 (19.6%) | 18,253 (24.2%) | 4,589 (28.6%) |
| ≥4 to <7 days | 4,477 (2.4%) | 2,347 (2.4%) | 3,654 (4.8%) | 1,533 (9.6%) |
| ≥7 to <14 days | 7,615 (4.0%) | 1,200 (1.3%) | 3,905 (5.2%) | 1,257 (7.2%) |
| ≥14 days | 10,982 (5.8%) | 907 (0.9%) | 3,911 (5.2%) | 1,129 (7.0%) |
| **Missing age at admission** | 1,277 | 721 | 522 | 54 |
| **Median (IQR)** | 0 (0-1) | 0 (0-0) | 0 (0-1) | 1 (0-3) |
| **Gestational age** | N= 160,924 | N=89,420 | N=66,064 | N=10,272 |
| **Extremely preterm (<28 weeks)** | 3,815 (2.4%) | 3,594 (4.0%) | 1,506 (2.3%) | 505 (4.9%) |
| **Very preterm (≥28 to <32 weeks)** | 11,915 (7.4%) | 8,595 (9.6%) | 5,551 (8.4%) | 1,558 (15.2%) |
| **Late preterm (≥32 to <37 weeks)** | 40,964 (25.5%) | 22,440 (25.1%) | 18,437 (27.9%) | 3,298 (32.1%) |
| **Term (≥37 weeks)** | 104,230 (64.8%) | 54,791 (61.3%) | 40,570 (61.4%) | 4,911 (47.8%) |
| **Missing gestational age** | 31,901 | 7,539 | 10,020 | 5,839 |
| **Mean (95%CI)** | 36.3 (36.25-36.28) | 36.3 (36.27-36.33) | 36.4 (36.37-36.42) | 35.3 (35.2-35.3) |
| **Birth weight** | N=178,737 | N= 95,944 | N= 75,475 | N=12,181 |
| **Extremely low birth weight <1000g** | 3,049 (1.7%) | 2,964 (3.1%) | 1,919 (2.5%) | 586 (4.8%) |
| **Low birth weight 1500-1999g** | 24,547 (13.7%) | 13,143 (13.7%) | 10,432 (13.8%) | 1,691 (13.9%) |
| **Low birth weight 2000-2499g** | 33,467 (18.7%) | 13,303 (13.9%) | 12,187 (16.2%) | 1,695 (13.9%) |
| **≥2500g** | 103,739 (58.0%) | 58,629 (61.1%) | 45,424 (60.2%) | 6,816 (56.0%) |
| **Missing birth weight** | 13,278 | 1,015 | 609 | 3,930 |
| **Mean (95%CI)** | 2540 (2537-2544) | 2647 (2641-2652) | 2630 (2624-2636) | 2518 (2501-2534) |
| **Length of stay** | N=186,260 | N=96,751 | N= 75,560 | N=16,037 |

|  |  |  |  |  |  |
| --- | --- | --- | --- | --- | --- |
|  | **<1 day** | 14,495 (7.8%) | 4,471 (4.6%) | 2,922 (3.9%) | 763 (4.8%) |
|  | **≥1 to <3 days** | 77,410 (41.6%) | 22,206 (23.0%) | 22,353 (29.6%) | 2,360 (14.7%) |
|  | **≥3 to <7 days** | 63,548 (34.1%) | 34,954 (36.1%) | 30,297 (40.1%) | 5,273 (32.9%) |
|  | **≥7 to <14 days** | 20,339 (10.9%) | 18,140 (18.8%) | 12,382 (16.4%) | 4,968 (31.0%) |
|  | **≥14 days** | 10,468 (5.6%) | 16,980 (17.6%) | 7,606 (10.1%) | 2,673 (16.7%) |
|  | **Missing** | 5,755 | 208 | 524 | 74 |
|  | **Median (IQR)** | 3 (1-5) days | 5 (2-9) days | 4 (2-7) days | 6 (3-10) days |
| **Sex** |  | N=181,846 | N=96,794 | N=75,877 | N=16,087 |
|  | **Male** | 99,962 (55.0%) | 54,371 (56.2%) | 41,619 (54.9%) | 9,016 (56.1%) |
|  | **Female** | 81,813 (45.0%) | 42,396 (43.8%) | 34,210 (45.1%) | 7,066 (43.9%) |
|  | **Indeterminant** | 71 (0.04%) | 27 (0.03%) | 48 (0.06%) | 5 (0.03%) |
|  | **Missing/not recorded/not readable** | 10,169 | 165 | 207 | 24 |
| **Place of birth** |  | N= 187,403 | N=90,259 | N= 75,827 | N=16,099 |
|  | **Inborn** | 146,538 (78.2%) | 75,672 (83.8%) | 49,516 (65.3%) | 8,145 (50.6%) |
|  | **Outborn** | 40,865 (21.8) | 14,587 (16.2) | 26,311 (34.7) | 7,954 (49.4) |
|  | **Missing/not recorded/not readable** | 4,612 | 6,700 | 257 | 12 |
| **Condition at discharge** |  | N=189,967 | N=96,876 | N= 75,734 | N=16,100 |
|  | **Alive** | 163,899 (86.3%) | 83,639 (86.3%) | 66,010 (87.2%) | 13,404 (83.3%) |
|  | **Dead** | 26,068 (13.7%) | 13,237 (13.7%) | 9,724 (12.8%) | 2,696 (16.8%) |
|  | **Missing/not recorded/not readable** | 2,048 | 83 | 350 | 11 |

### Service readiness

Across 64 hospitals in Malawi, Kenya, Tanzania, and Nigeria, bed occupancy showed marked within- and between-country variability. Overall median occupancy across hospital-months was 65% (IQR: 38%–106%), with 28% exceeding 100%. At hospital-level, the median of monthly medians ranged from 14% to 205%; 20 hospitals (31%) recorded >100% occupancy in more than half of months (**Figure 2**). Overcrowding was most consistent in Malawi and Kenya, where 39% of Kenyan hospitals exceeded capacity in most months. No hospital in Nigeria or Tanzania met this threshold.

**Figure 2.**
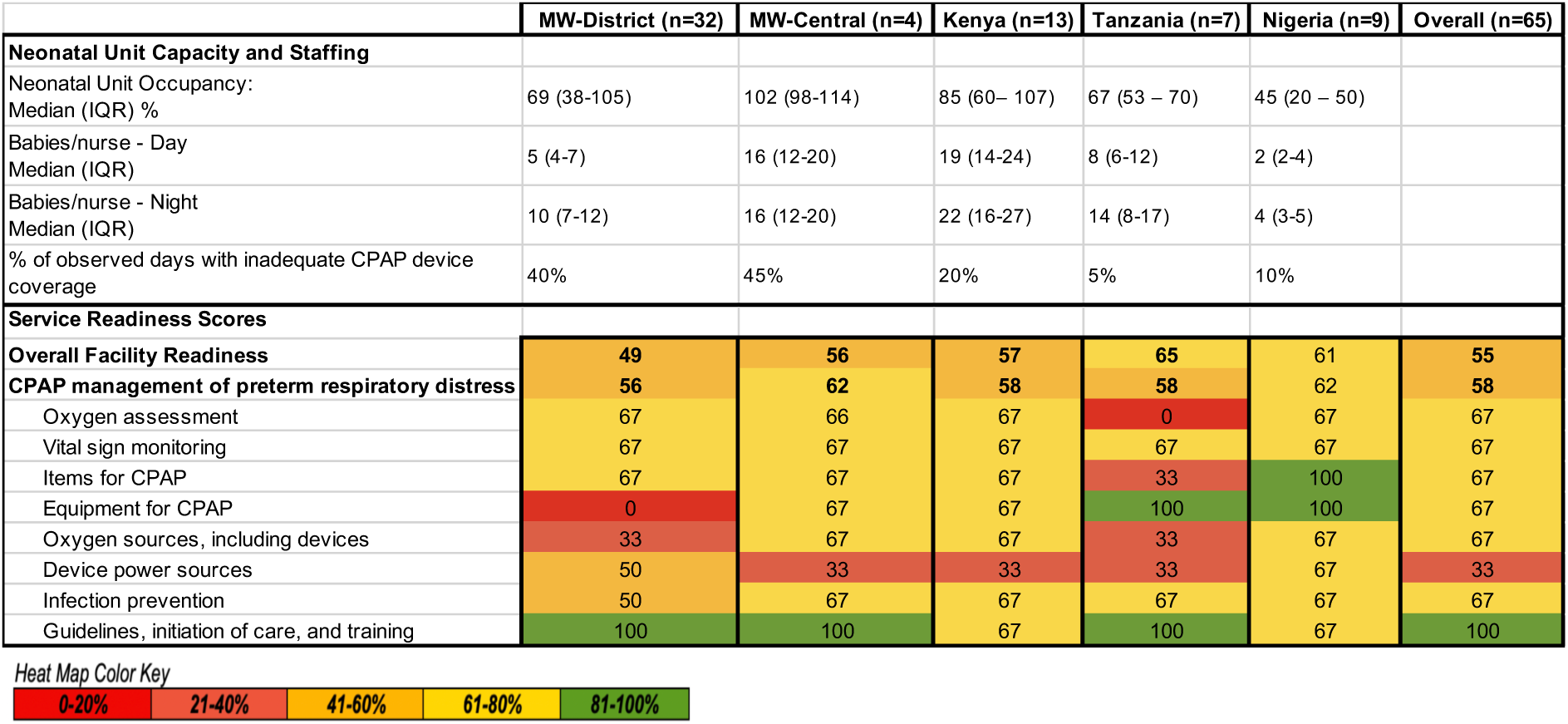
Neonatal respiratory service availability (April 2024 - March 2025) and respiratory service readiness scores (2023) across implementing hospitals. Respiratory service availability calculated from weekly hospital context-tracker data (**Additional File 9**). Staffing ratios described as babies/nurse for day and night shifts. No facility data available for one hospital in Malawi. IQR=interquartile range, %=percentage. Heat map of median health services readiness scores for each indicator related to CPAP from 2023 cross-sectional HFA assessment data. Colour key depicts percentage ranges for each indicator. Adapted from Penzias et al [40], with link to HFA Tool in **Additional File 8**. No facility data available for one hospital in Malawi. *MW=Malawi; CPAP=continuous positive airway pressure.

Between April 2024 and March 2025, 18,621 hospital-days were assessed for CPAP device availability relative to estimated daily need. Nearly one-third (31%) of days reflected inadequate coverage, defined as ≥1.5 eligible babies per device. Malawi had the greatest burden, with 43% of 10,962 days above this threshold (45% central; 40% district hospitals), followed by Kenya (20% of 4,073 days). Tanzania and Nigeria had fewer days of inadequate coverage (10% and 5%, respectively), with median ratios of 0. At hospital level, the median percentage of inadequate-coverage days was 16% (IQR: 4%–47%).

Across hospitals, the median babies per nurse was 6 (IQR: 4–12) during day shifts and 11 (IQR: 7–18) at night (**Figure 2**), with substantial variability. Staffing coverage was most inadequate in Kenya, particularly at night, and least inadequate in Nigeria.

HFA service readiness scoring illustrated gaps in power sources for devices in Malawi’s central hospitals, and in facilities in Kenya and Tanzania **(Figure 2)**. Guidelines, initiation of care, and training showed higher scores (67-100%) across hospitals in all countries.

### CPAP coverage

Neonates eligible to receive CPAP were first identified: neonates with a birthweight <1500g, neonates with respiratory symptoms or hypoxia weighing between 1500-1999g, and any neonates with a clinical diagnosis of RDS above these weights, yielding a total of 70,083 eligible neonates (**Figure 1**).

Overall CPAP coverage across eligibility groups and countries was described in **Additional File 3**. Missingness (defined as true missing, not recorded, or not readable) in the CPAP administration data was noted to be highest in Tanzania, with 22% missingness, followed by 12% missingness in Nigeria, mostly due to a lack of clinical chart documentation (99% of missing records), with certain facilities contributing most to missingness **(Additional File 4)**. Complete case analysis gave a pooled CPAP coverage proportion of 34% (95% CI 33.6-34.3%), with a total of 22,006 CPAP-eligible neonates receiving CPAP out of 64,761 CPAP-eligible neonates with non-missing CPAP data across all admissions. Pooled coverage was highest for newborns weighing 1000-1499g (43%, 95% CI 42.6-43.8%), and lowest for newborns with RDS and birth weight 2000g or above (16%, 95% CI 15.0-16.3%), though variability existed between countries. CPAP coverage was highest in Nigeria for all eligibility groups **(Additional File 5)**, though Nigeria had lower admission numbers than other countries **(Table 2)**.

CPAP coverage over time was assessed using mean coverage proportions at neonatal unit-level, with pooling of means of means at country-level, over quarterly time intervals **(Figure 3)**. In Malawi’s central hospitals, CPAP coverage increased from a mean of 14% (95% CI -1 to 30%) to 38% (95% CI 14-62%) from 2021 quarter 1 to 2024 quarter 4, a 24% increase (p<0.05). In Malawi’s district hospitals, coverage increased from a mean of 13% (95% CI 9-17%) to 44% (95% CI 37-52%), representing a 31% increase (p<0.0001).

**Figure 3.**
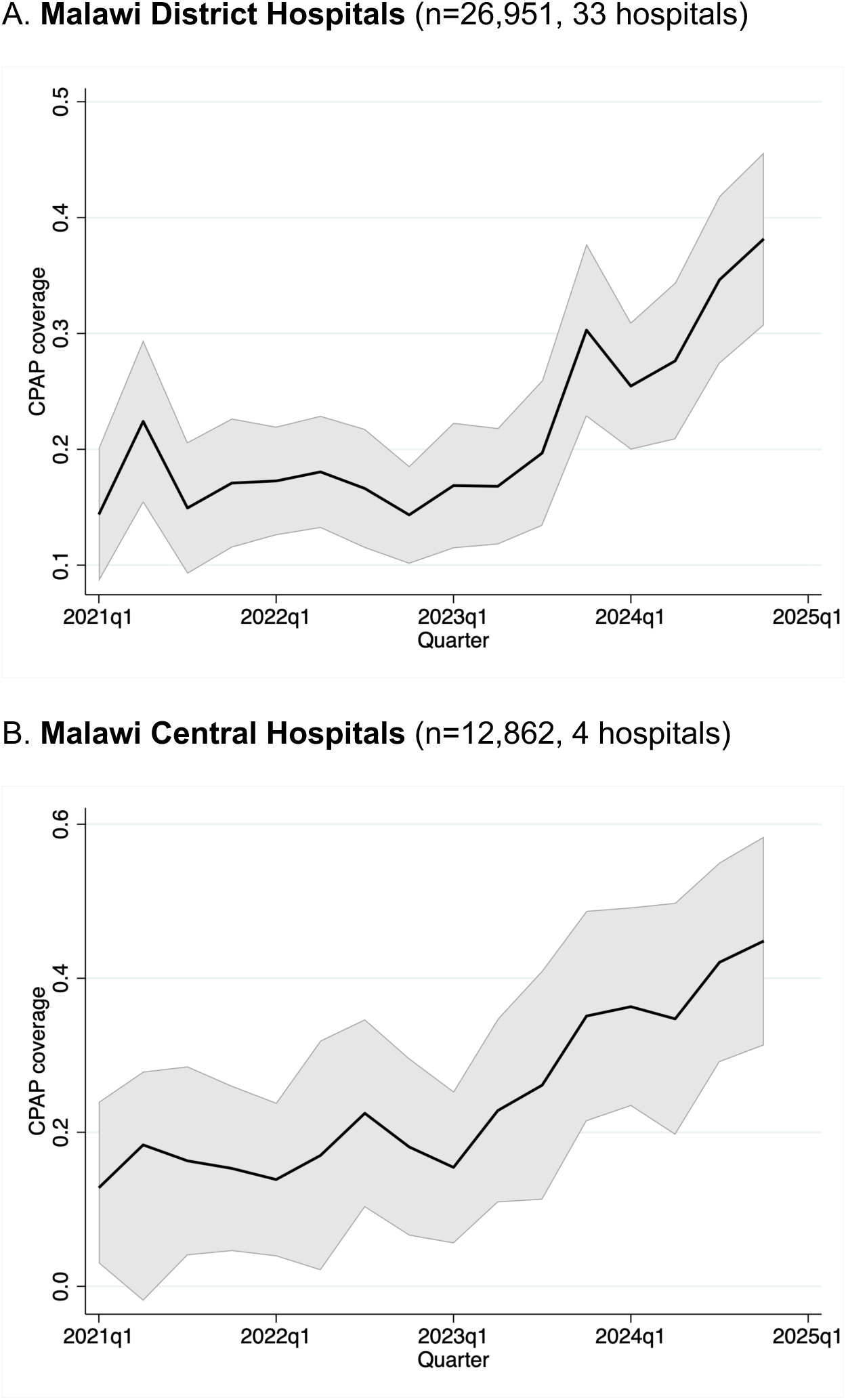
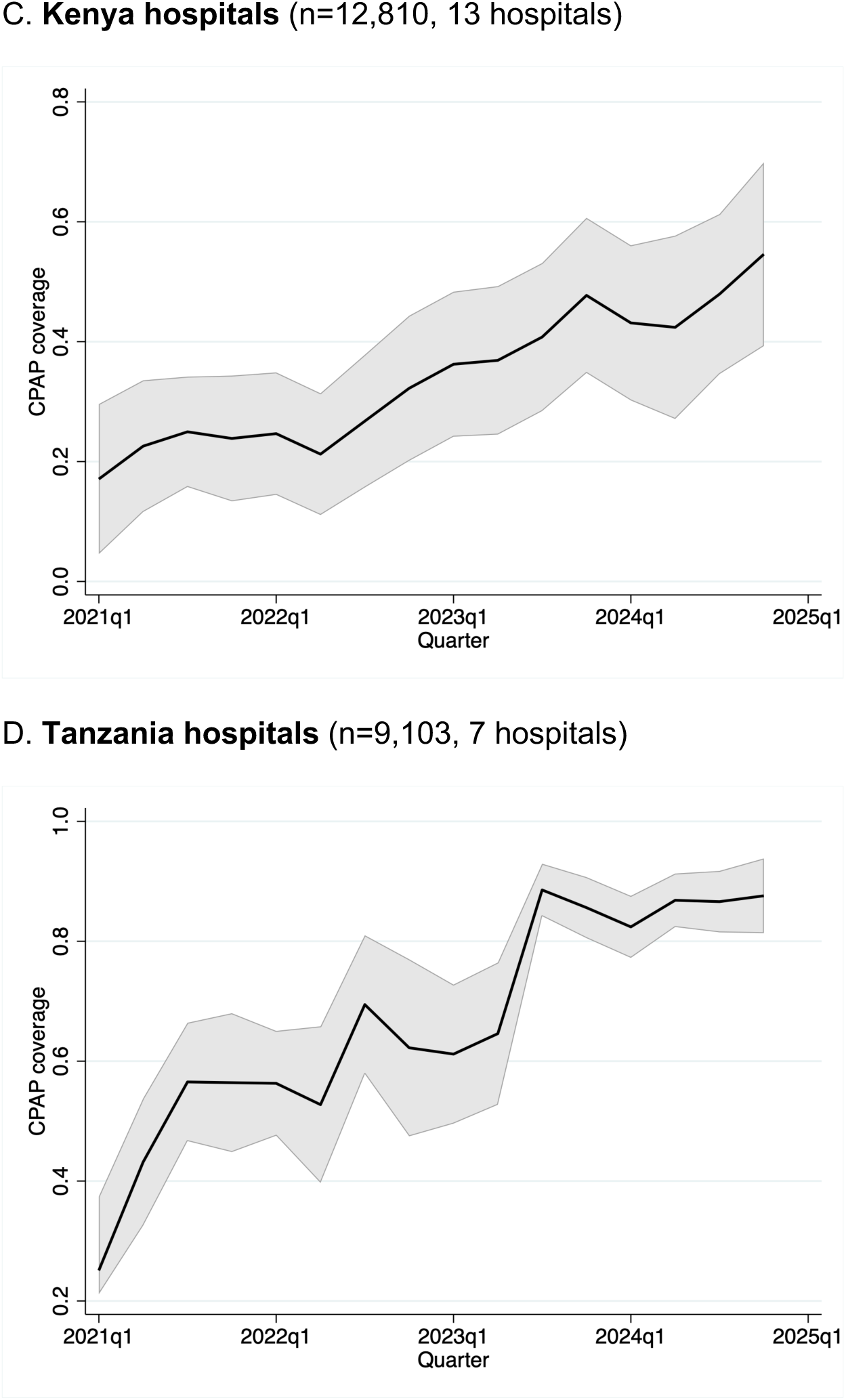
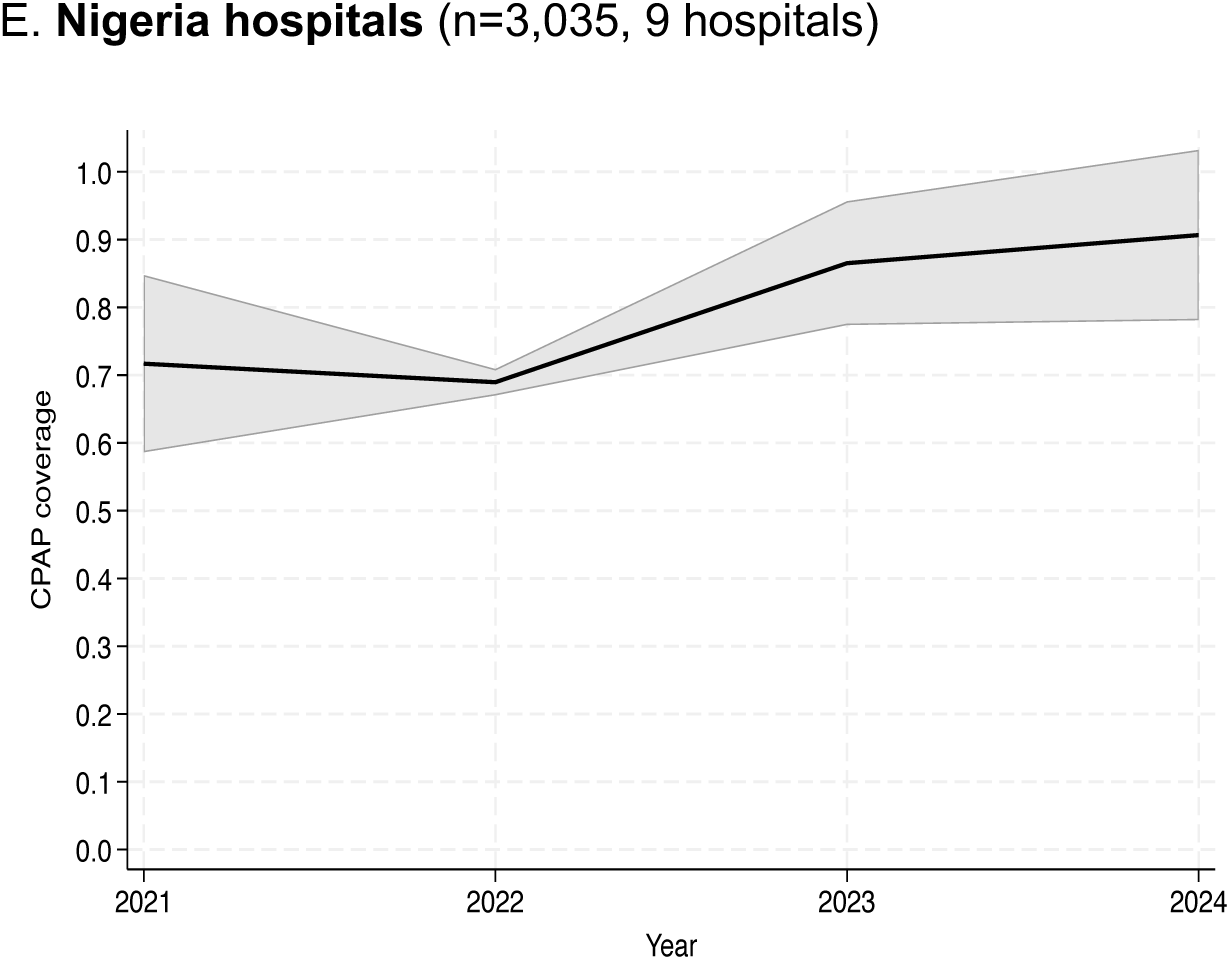
Mean coverage proportions for CPAP-eligible neonates using hospital-level analysis per quarter for each country (January 2021 - December 2024). Country-level mean coverage proportions obtained from hospital-level mean coverage proportions in each country. Mean coverage plotted over quarterly time periods, except for Nigeria, where coverage plotted annually. 95% confidence intervals depicted in grey shading around mean coverage lean. Total n= 64,761 CPAP-eligible neonates; Malawi central hospitals (n=12,862), Malawi district hospitals (n=26,951), Kenya hospitals (n=12,810), Tanzania hospitals (n=9,103), Nigeria hospitals (n=3,035).

Kenya’s mean CPAP coverage increased 38% (p=0.001) from a mean of 17% (95% CI 4-30%) in 2021 quarter 1 to 55% (95% CI 43-66%) in 2024 quarter 4. Tanzania achieved the highest increase in coverage, 63% increase in mean coverage over the analysis period (p=0.0001), increasing from an initial mean coverage of 25% (95% CI 3-48%) to a mean of 88% (95% CI 76-99%).

Nigeria demonstrated coverage means ranging from 79% (95% CI 40-118%) to 91% (95% CI 75-106%) across the analysis period but had wide, overlapping confidence intervals, and a lower number of admissions compared to other countries. However, there were non-overlapping intervals in 2022 and 2024 suggesting an increase in coverage between those years.

### Quality cascades

CPAP quality of care was evaluated for each country with clinical cascades for CPAP-eligible neonates who received CPAP during admission. The first cascade focused on timing of CPAP initiation and duration of treatment for the first episode of CPAP administration **(Figure 4A)**, as most babies (95%) with data who were managed with CPAP received one episode. Kenyan facilities were omitted from this analysis given CPAP timing data were not available for much of the analysis period. Across the pooled dataset of all CPAP-eligible neonates, the median timing for CPAP initiation was 8 hours of life (IQR 2-26 hours) and median CPAP duration was 51 hours (IQR 24-103 hours). Forty-two percent (95%CI 41-43%) of all CPAP-eligible neonates who received CPAP with associated timing data started within the first day of life (DOL) and continued for more than 24 hours.

**Figure 4.**
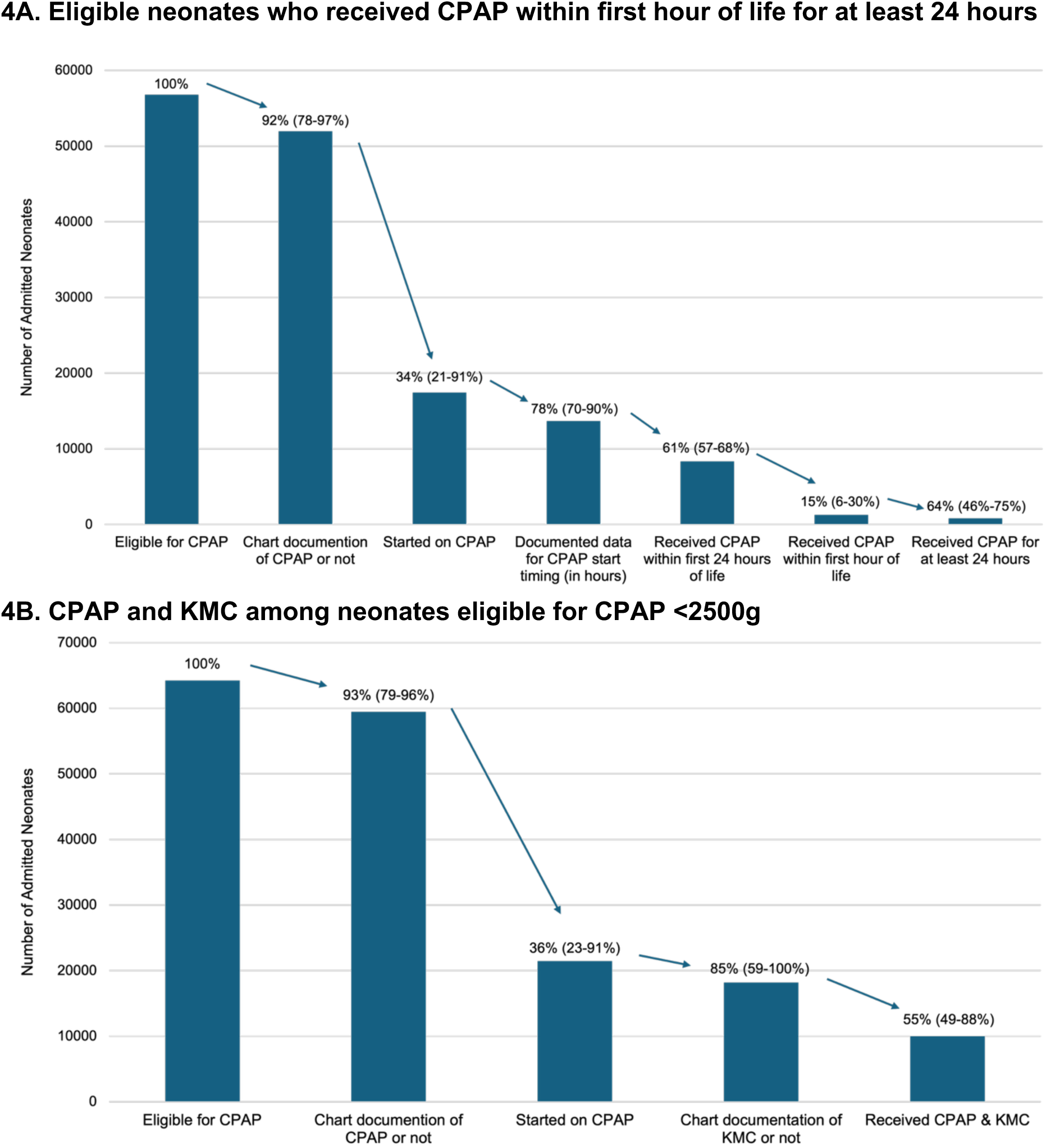
Quality of Care Cascades for Neonates Receiving CPAP A. Clinical quality cascade reflecting timing of CPAP initiation and duration of therapy, pooled across all countries (January 2021-December 2024). Initial n= 56,770. In this cascade, the first indicator is the number of CPAP-eligible neonates and constitutes 100%. This number then serves as the denominator for the subsequent indicator in the cascade. Each subsequent indicator uses the preceding total as its denominator. Results were pooled across Malawi central and district hospitals, Tanzanian hospitals, and Nigerian hospitals, with the inter-country range illustrated in parentheses next to overall percentages above bars on the graph. Kenyan data excluded. **B. Clinical quality cascade reflecting CPAP and KMC co-administration, pooled across all countries (January 2021-December 2024).** Initial n = 64,234 neonates <2500g. Results were pooled across all four countries, with the inter-country range illustrated in parentheses next to overall percentages above bars on the graph. See **Additional File 6** for additional cascades.

Pooled CPAP coverage was 33% overall, ranging from 21% of eligible newborns in Malawian district hospitals to 91% in Nigeria **(Figure 4A)**. Of these neonates started on CPAP, documentation of timing was high across all three countries. Initiation within 24 hours of life ranged from 57% of babies receiving CPAP in Tanzania to 67% of those in Malawian central hospitals. Between 6% of newborns in Malawian central hospitals and 30% in Nigerian hospitals were initiated within the first HOL. Between 46% of newborns admitted in Nigeria and 75% of those admitted in Malawian central hospitals continued CPAP for at least 24 hours. For neonates weighing <1500g and hence eligible for prophylactic CPAP, the pooled CPAP coverage proportion was higher at 46%, with Nigeria performing highest at 93% **(Additional File 6A).** Only 10% of these neonates who started CPAP with documentation of all indicators were initiated on early CPAP during their first HOL. The proportion was lowest in Malawi’s central hospitals (5%) and again highest in Nigerian units (20%).

CPAP quality was assessed in terms of co-administration with another high-impact intervention, kangaroo mother care (KMC), across all four countries **(Figure 4B).** KMC co-administration among newborns receiving CPAP with documentation on indicators ranged from 48% in Tanzanian hospitals to 88% in Nigerian hospitals.

To further understand quality, we assessed CPAP device types in Malawi, Tanzania, and Nigeria from June 2024 through December 2024 **(Figure 5).** Tanzania had highest proportions of CPAP devices using humidified air only or heated humidified air (70% of its devices). Nigeria had the highest proportion of improvised CPAP devices (22%). Device data were not available for Kenya.

**Figure 5.**
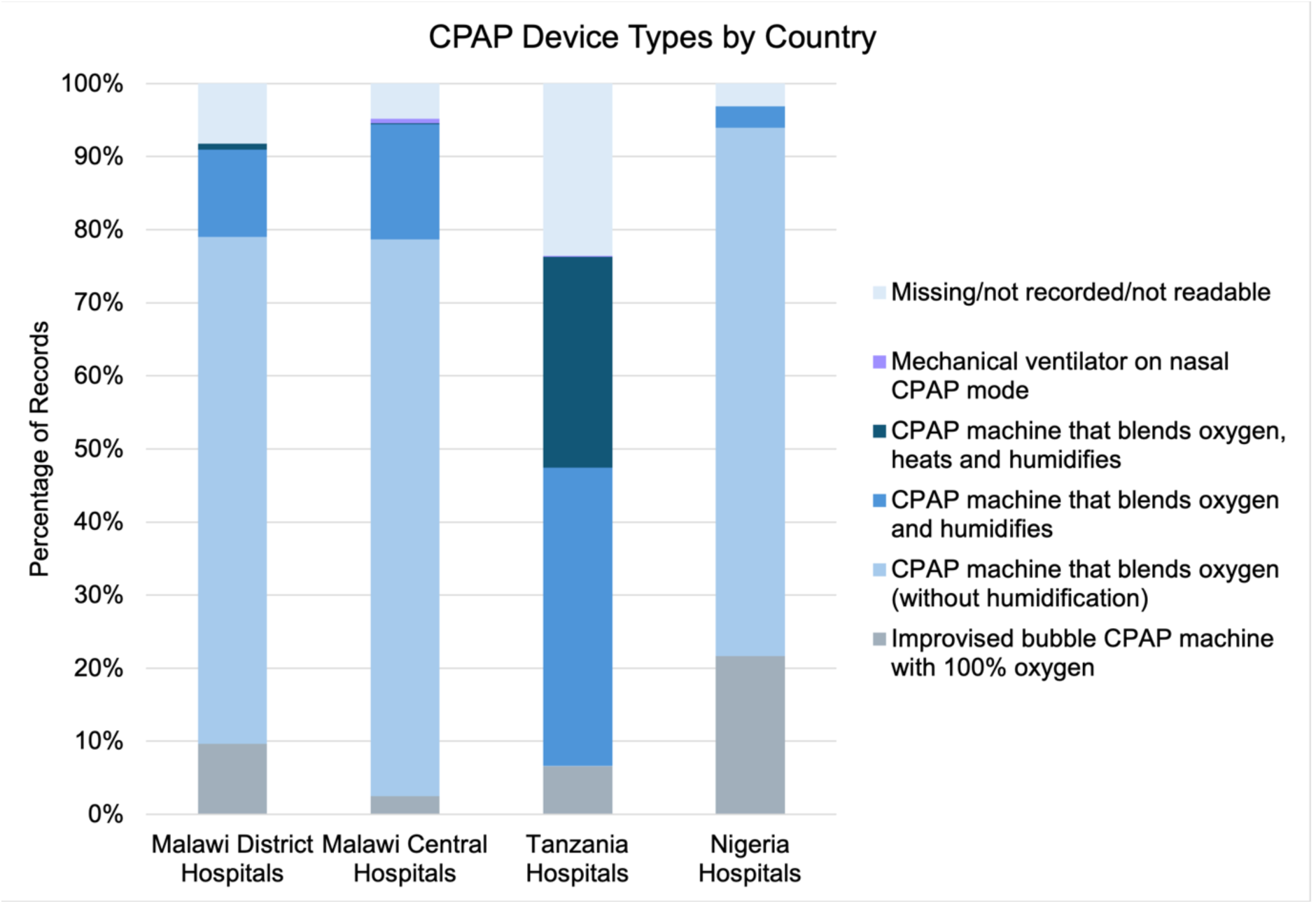
Types of CPAP devices used in each country’s hospitals for CPAP-eligible neonates (June 2024 - December 2024). The legend depicts the different CPAP device options available across implementing hospitals and associated colours. N=4,020 neonates with available data. Malawi district hospitals (n=1767), Malawi central hospitals (n=681), Tanzania hospitals (n=1,157), Nigeria hospitals (n=415).

### Outcomes

A clinical severity score comprising gestational age, oxygen saturation on admission, and hypothermia on admission was developed for survival modelling, with low (3-4), moderate (5-6), and a high (7-9) severity score groupings. Predicted probability of surviving to discharge was modelled among CPAP-eligible neonates <2500g **(Figure 6)**. The mixed-effects logistic regression model demonstrated good discrimination for survival with an AUC of 0.74, **(Figure 6C)**. After adjustment for covariates and hospital-level clustering, the association between CPAP and survival differed significantly by clinical severity group **(Figure 6A, 6B)**. Newborns with high clinical severity treated with CPAP had a higher probability of survival (32%, 95% CI 29–36%) than newborns with high clinical severity who did not receive CPAP (23%, 95% CI 21–26%). CPAP-eligible newborns who did not receive CPAP had higher adjusted survival probabilities among low and moderate severity groups.

**Figure 6.**
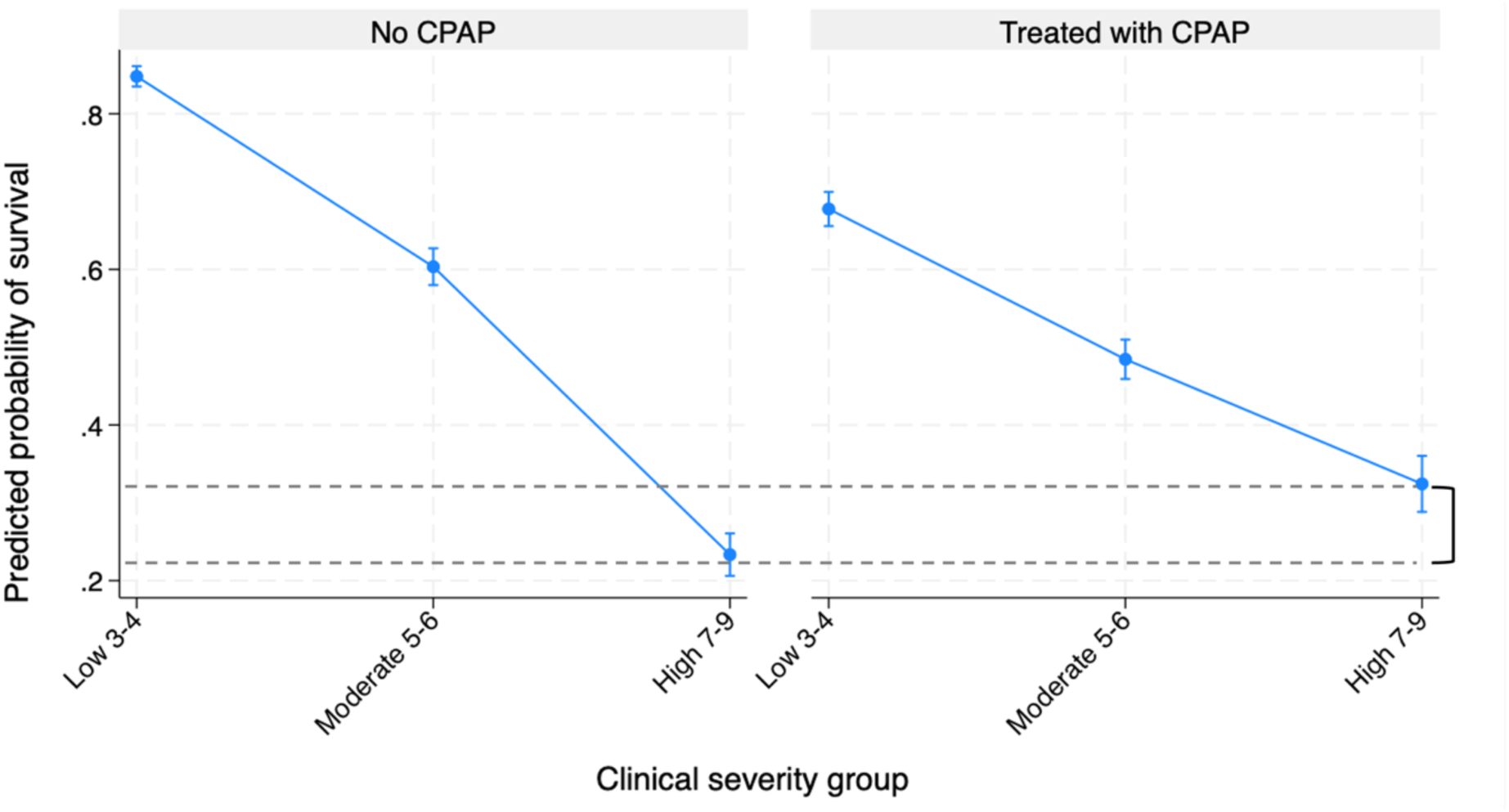
A. Predicted probability of survival among CPAP-eligible neonates <2500g. 95% Confidence intervals depicted by bars on plot. Bracket shows difference between predicted probabilities of survival between high severity groups with and without CPAP.

**Figure 6.**
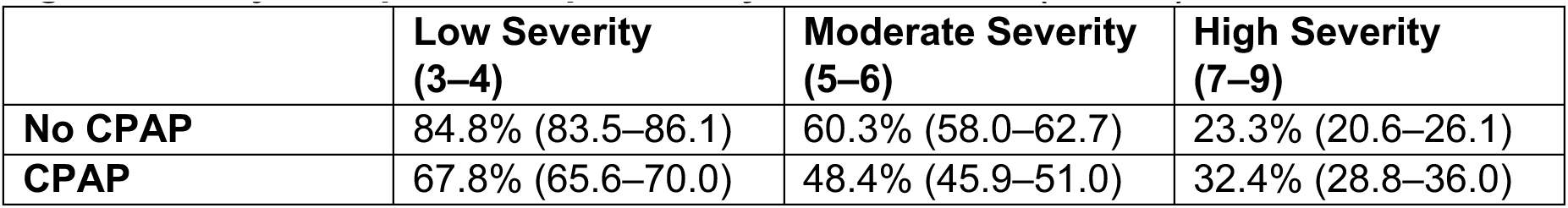
B. Adjusted predicted probability of survival, % (95% CI)

**Figure 6.**
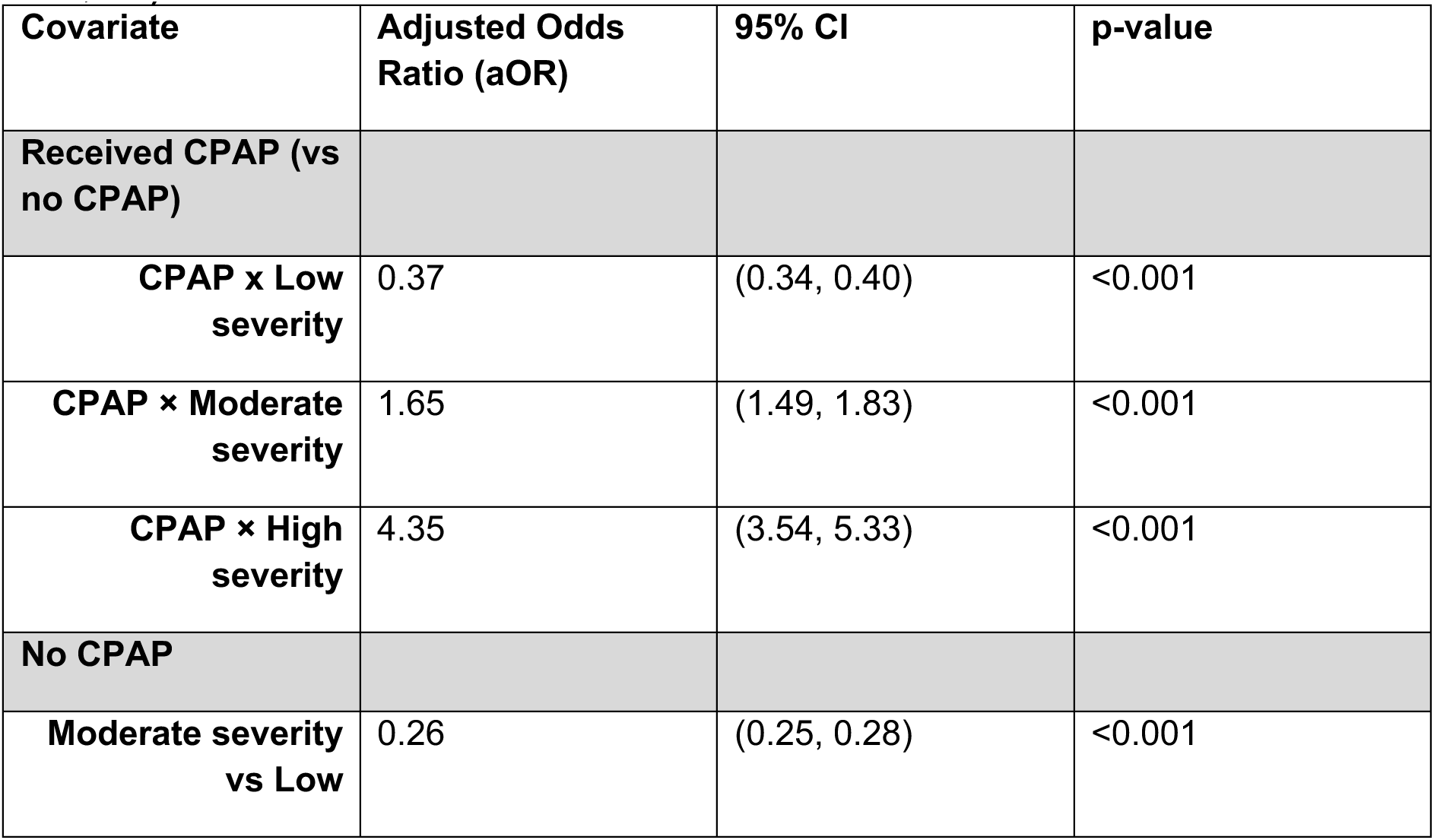
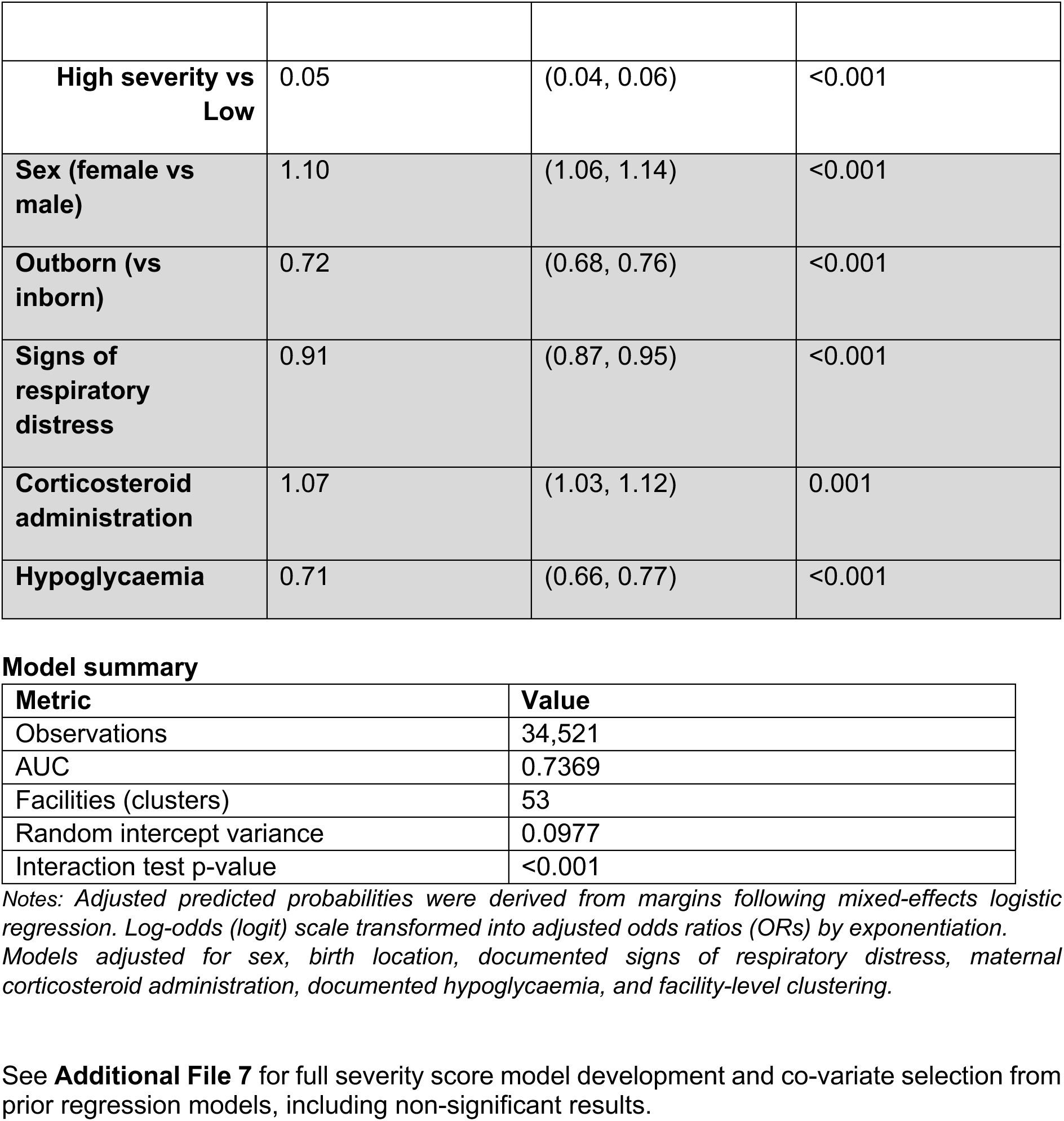
C. Adjusted associations for covariates (mixed-effects logistic regression, n=34,521)

Higher adjusted odds of survival were observed for female newborns (aOR 1.10, p<0.001) and for newborns whose mothers had received antenatal corticosteroids (aOR 1.07, p=0.001) **(Figure 6C)**. Probability of survival decreased with increasing clinical severity score. Lower adjusted odds of survival were seen for newborns with documented hypoglycaemia (aOR 0.71, p<0.001), documented signs of respiratory distress (aOR 0.91, p<0.001), and outborn newborns (aOR 0.72, p<0.001).

## DISCUSSION

Our large dataset included 375,255 neonatal admissions across 66 neonatal units in Malawi, Kenya, Tanzania, and Nigeria. This analysis provides a unique multi-country assessment of CPAP coverage, quality, and outcomes for neonates. Although CPAP is the recommended standard-of-care for the management of RDS in premature neonates, coverage and quality varied across implementing hospitals in this analysis. Increases in CPAP coverage in three countries is an encouraging finding. Our analysis demonstrated the life-saving effects of CPAP for newborns with severe illness. Gaps remain in both coverage and quality of care for CPAP including timely initiation, duration and CPAP device type.

Neonatal units across the four countries showed compounding structural constraints on the delivery of small and sick newborn care. Persistent bed overcrowding signals a chronic demand-supply mismatch that strains the full scope of unit function, from infection control to individualized clinical attention. Staffing shortages reinforced this picture, consistent with prior assessments of the same hospitals [41]. These workloads exceed thresholds at which nurses can reliably monitor, respond to deterioration, and deliver the range of interventions that SSNC requires.

CPAP device shortfalls are best understood as one expression of this broader resource-constrained environment rather than an isolated supply problem. HFA readiness scoring pointed to power infrastructure as the most critical unresolved gap in Malawi, Kenya, and Tanzania, while performance on guidelines, care initiation, and training was comparatively stronger across all countries - suggesting that soft systems gains have been made even where physical infrastructure remains fragile.

Despite illustrated gaps in hospital-level service readiness for CPAP, these were not predictive of actual coverage or impact, as there was clear improvement in coverage despite limitations. Literature often highlights measuring effective coverage as a function of contact point, service readiness, and outcome data [60]; but this analysis also highlights the need to assess quality of care. There is nonetheless a complex interplay between these components, as inadequate staffing levels may be a barrier to providing high-quality CPAP, even when devices are available.

### CPAP Coverage

CPAP coverage increased across each country’s hospitals during the analysis period (January 2021-December 2024). Tanzania had the largest increase in CPAP coverage overall, 63% (from 25% to 88%), followed by a 38% increase in Kenya (17% to 55%), 31% in Malawian district hospitals (13% to 44%), and 23% in Malawian central hospitals (14% to 38%). Nigeria had high baseline CPAP coverage (79%, 95% CI 40-118%) with lower admission numbers, hence increases were not statistically significant.

Coverage varied by birthweight group, though these were assessed in a pooled analysis rather than over time **(Additional File 3)**. The highest pooled coverage proportion (43%) was for babies with birth weights 1000-1499g, consistent with clear programmatic guidance on prophylactic CPAP for such neonates (33). Lower coverage (39%) for newborns <1000G is likely related to use of CPAP devices specifically licensed for neonates >1000g [23]; and clinical prioritisation of CPAP devices for newborns with higher birth weights, given well-described poorer outcomes in ELBW neonates in LMIC settings [61]. Symptomatic newborns ≥1500 grams had even lower coverage (31% in 1500-1999g group), possibly related to delays in recognising symptoms, or poorer prognosis once clinical illness was recognised, leading to prioritisation of other neonates. Misclassification of hypotonia for hypoxic ischemic encephalopathy (HIE) could also lead to lower coverage in higher weight groups [62, 63]. Clinician knowledge and training should accompany unit guidelines to avoid misclassification.

### CPAP Quality

Despite encouraging increases in CPAP coverage, quality indicators showed variability. In Tanzania, where the increase in coverage was highest, CPAP device types provided better care quality with higher proportions of humidified, as well as heated and humidified, blended oxygen systems [64]. However, a quality-of-care gap was noted in timing in Tanzanian hospitals, with the lowest proportion of eligible newborns initiating CPAP within the first day of life (57%). In Nigeria, high CPAP coverage did not coincide with high-quality CPAP device types, given the highest proportion of improvised CPAP devices were in Nigeria (22%). Care quality in Nigeria was higher for other indicators, with high co-administration of IV fluids, and higher coverage of early CPAP (30% within first HOL) compared to other countries’ hospitals.

Novel pooled CPAP quality of care cascades illustrated several actionable care quality indicators. A gap in CPAP quality around timely initiation showed 61% of neonates with timing data received CPAP on their first day of life, and even fewer within one HOL (15%). This initiation gap may lead to poorer outcomes [30], especially among infants who may look well at first, particularly in the 1500-1999g category where symptom and hypoxia recognition are key to initiating CPAP [44]. The gap for early prophylactic initiation within the first HOL among newborns weighing <1500g (10%) could reflect delays in the delivery room setting [65], which is a critical part of the neonatal care pathway. Multidisciplinary team coordination between the labour and delivery, and neonatal departments, may enable earlier CPAP [66], though this also depends on availability of transportable CPAP.

KMC co-administration rates also demonstrated a gap, with 55% of neonates on CPAP with available data receiving both interventions. There was an accompanying data quality gap, a lack of detailed timing of KMC, so it was not possible to investigate simultaneous co-administration. KMC and CPAP co-administration gaps could be explained by many issues, such as caregiver and staff education and comfort around maintaining CPAP therapy, with its associated accessories and monitoring procedures [44], as noted for other interventions like intravenous fluids [67]. Unit design and resources are also important for enabling KMC [68]. Neonates on CPAP are likely to be less clinically stable and may have additional complications making KMC more challenging. These findings underscore the need for improved unit design to facilitate safe CPAP-KMC integration [68] as well as staff training on effective KMC before stabilisation, as immediate KMC is key for improving outcomes [69].

### Outcomes

Predicted probability of survival correlated well with clinical severity across CPAP-eligible neonates <2500g. High clinical severity leads to death without CPAP (predicted probability of survival 23%, 95% CI 21–26%), but survival increases with CPAP (predicted probability of survival 32%, 95% CI 29–36%). This positive association with CPAP was not observed in low and moderate severity groups. This likely reflects confounding wherein neonates who appeared clinically worse despite similar severity scores had escalation of care to CPAP. It also likely reflects unmeasured acuity not accounted for in our clinical scoring model, as variable selection was limited by factors such as data availability and circularity with eligibility criteria for the intervention.

Newborns whose mothers had received antenatal corticosteroids had higher odds of survival, consistent with improved lung maturation and lower risk of RDS [59]. This points to an important opportunity for preventive maternal care when preterm births are anticipated [70]. As expected, odds of survival were poorer for newborns with documented signs of respiratory distress, consistent with illness severity. Hypoglycaemia was also associated with lower odds of survival, consistent with other studies [58], which points to the importance of quality comprehensive preterm care, which does not only address respiratory care. Interventions like KMC which improve early breastfeeding are critical for quality care in CPAP-eligible newborns [69] and are an integral part of the bundle of care for preterm.

Outborn infants represent a high proportion of admissions (**Table 2**) and had lower odds of survival. Increased neonatal mortality in South Africa and Nigeria for outborn infants has been well-described [71–73], leading to efforts to increase facility deliveries and strengthen referral pathways. Referral delays are also of major concern in high-burden settings, leading to adverse outcomes [74, 75]. Strengthening referral pathways must be prioritized to improve care [76].

### Strengths and Limitations

Strengths of this analysis include the large multi-country neonatal inpatient dataset (over 375,000 newborn admissions), with robust standardised data (NID) linked to routine health systems and systematic data quality checks. Another strength is novel conceptualisation to quantify quality using individual data in clinical quality of care cascades, learned from HIV clinical cascades (35,36). Admitted newborns to NEST360-implementing units are likely to share characteristics with other admitted newborns in the region, and parallels may be drawn to improve care quality in other hospitals in the region.

The severity score constructed to predict survival is also an important addition to the literature on newborn outcomes [10] and is in the process of analysis in another dataset.

Limitations include missing data, which may have led to measurement bias for more detailed information regarding CPAP use particularly with missingness varying along the cascades and higher in Tanzania and Nigeria (22% and 12% of administration data, respectively). Most hospitals implementing with NEST360 have data linked to routine government systems [37], but Kenya used data from a larger research database [38] which had gaps for some key QI variables. As this analysis used routinely collected data, some important facets of care quality were not assessed.

While this analysis focused on CPAP, other interventions in the respiratory care package were not included, such as management for apnoea of prematurity. Though antenatal corticosteroids were assessed as a covariate, more antenatal data on RDS prevention would be useful in forthcoming analyses. Future research should examine coverage and care quality for the entire respiratory care bundle, and maternal-newborn linkages for high-quality care.

## Conclusion

There has been significant improvement in CPAP coverage over time across the NEST360-implementing hospitals, but quality of care remains critical to drive progress for better management and outcomes for preterm neonates. To achieve high-quality CPAP, neonatal units must utilise high-quality and context-appropriate CPAP devices at the right time and duration for the right babies. Other high-impact newborn interventions like KMC will also lead to optimised care and outcomes [19, 69, 77], as will managing co-morbid conditions like hypothermia and hypoglycaemia, and optimising antenatal corticosteroid use. Our global community must continue to work toward achieving SDG 3.2 for vulnerable preterm newborns by closing CPAP coverage and care quality gaps through resource allocation, trained newborn clinical staff, and use of data for action. Providing high-quality respiratory care for every small and sick newborn who needs this care everywhere is necessary for faster progress on national and global targets.

## Availability of data and materials

All collaborating partners in the NEST360 Alliance jointly developed and signed data sharing and transfer agreements. The NID and HFA tools, data dictionaries and training are available from NEST360/UNICEF Implementation Toolkit for Small and Sick Newborn Care [42] and in **Additional Files 8,9**.

## Abbreviations

aOR: Adjusted odds ratio
CI: Confidence interval
CIN: Clinical Information Network
CPAP: Continuous positive airway pressure
DOL: Day of life
ELBW: Extremely low birth weight
G: Grams
HFA: Health Facility Assessment
HIC: High income country
HIE: Hypoxic ischemic encephalopathy
HSBB: Health service building block
HOL: Hour of life
I: Intravenous
KMC: Kangaroo mother care
LBW: Low birth weight
LMICs: Low- and middle-income countries
LOS: Length of stay
MW: Malawi
NEST360: Newborn Essential Solutions and Technologies
NID: Newborn Inpatient Dataset
NMR: Neonatal mortality rate
OR: Odds ratio
QI: Quality improvement
RDS: Respiratory distress syndrome
SDG: Sustainable development goal
SpO2: Oxygen saturation
STROBE: Strengthening the Reporting of Observational Studies in Epidemiology
VLBW: Very low birth weight
WHO: World Health Organization

## Data Availability

All collaborating partners in the NEST360 Alliance jointly developed and signed data sharing and transfer agreements. The NID and HFA tools, data dictionaries and training are available online from NEST360/UNICEF Implementation Toolkit for Small and Sick Newborn Care and in Additional Files 8,9.

https://newborntoolkit.org/toolkit/data-systems-qi

## Acknowledgements

We thank all the newborns and their mothers whose data were included in this analysis. We also acknowledge all hospital teams implementing with NEST360 and the teams involved as part of the *Newborn Inpatient Dataset Group* and *Hospital Facility Assessment Group,* and all those collecting and using data for action. Finally, we are grateful to fellow researchers and guest editors who reviewed this paper, and for the input from managing editors at BMC and within NEST360 including Caroline Noxon, Kristina Shemwell, Harriet Ruysen, William M. Macharia, and Joy E. Lawn.

## Declarations

### Author Information

Kristina Shemwell and John Wainaina Joint first authors.

James H. Cross, Kondwani Kawaza & Elizabeth M. Molyneux Joint senior authors.

### Collaborative groups

#### NEST360 Neonatal Inpatient Dataset and Data Systems Collaborative Group and Context Tracker

James H. Cross; Christine A. Bohne; Lucas Malla; Morris Ondieki Ogero; John Wainaina; Julius Thomas; Eric O. Ohuma; Samuel K. Ngwala; Joseph Misyenje; Msandeni Chiume; Josephine Shabani; Irabi Kassim; Prosper Mshana; Jacqueline Minja; Honorati Masanja; Franklin Okech; Vincent Ochieng; Olabisi O. Dosunmu; Rebecca E. Penzias; Kristina Shemwell; Maria Oden; Rebecca Richards-Kortum; Joy E. Lawn.

#### NEST360 Health Facility Assessment Collaborative Group

Rebecca E. Penzias; Christine A. Bohne, Morris Ondieki Ogero, Samuel K. Ngwala; Evelyn Zimba; Msandeni Chiume; Joseph Misyenje; George Banda; Lucas Malla, Eric O. Ohuma; Josephine Shabani; Irabi Kassim; Prosper Mshana; Jacqueline Minja; Robert Tillya; Vincent Ochieng; George Okello; David Gathara; Olabisi O. Dosunmu; Opeyemi Odedere; James H. Cross; Edith Gicheha; Elizabeth M. Molyneux, Maria Oden, Rebecca Richards-Kortum; Joy E. Lawn.

### Author Contributions

This analysis was done as part of ongoing work with the NEST360 Alliance and partners. Those involved in this paper are named in the collaborative groups and recognised for their role in data collection, management, analysis, and manuscript review. The NEST360 Complex Evaluation was conceptualised by the NEST360 Alliance Team, facilitated by JEL. All collaborators contributed to the design of the study protocol. KS, JEL and JHC developed the detailed research questions and overall analysis plan for this paper. These were refined with inputs from co-authors and the wider NEST360 teams. Analysis was undertaken by KS and JW, who also drafted the manuscript. JEL, CB, JHC, KK, EMM, provided valuable guidance and support on analyses and manuscript development. REP assisted with HFA data management and statistical analysis. KS and JEL developed clinical severity score in consultation with LM and wider team. All authors reviewed and helped to revise the manuscript. All authors reviewed and agreed on the final version. The authors’ views are their own, and not necessarily from any of the institutions they represent.

**Corresponding author** Correspondence to Kristina Shemwell.

## Ethics declarations

### Ethics approval and consent to participate

Ethical approval was received in each country from respective ethical review committees **(Additional File 2)** and the London School of Hygiene & Tropical Medicine ethics committee (no. 30612). The NEST360 Alliance data sharing agreement covered data sharing between sites. Where applicable, national data transfer agreements were acquired. No individual consent was required for the study as data were collected as part of routine data collection at each site.

### Consent for publication

Not applicable.

## References

[1] L. Liu et al., “Global, regional, and national causes of under-5 mortality in 2000-15: an updated systematic analysis with implications for the Sustainable Development Goals,” (in eng), Lancet, vol. 388, no. 10063, pp. 3027–3035, Dec 17 2016, doi: 10.1016/s0140-6736(16)31593-8.

[2] UN-IGME, “Levels & trends in child mortality child mortality report,” 2024. [Online]. Available: https://data.unicef.org/resources/levels-and-trends-in-child-mortality-2024/

[3] World Health Organization. “Newborn mortality.” https://www.who.int/news-room/fact-sheets/detail/newborn-mortality (accessed 1 June, 2025).

[4] UNICEF. “Neonatal Mortality.” https://data.unicef.org/topic/child-survival/neonatal-mortality/ (accessed 1 June, 2025).

[5] L. Hug, M. Alexander, D. You, and L. Alkema, “National, regional, and global levels and trends in neonatal mortality between 1990 and 2017, with scenario-based projections to 2030: a systematic analysis,” (in eng), Lancet Glob Health, vol. 7, no. 6, pp. e710–e720, Jun 2019, doi: 10.1016/s2214-109x(19)30163-9.

[6] H. Blencowe et al., “National, regional, and worldwide estimates of preterm birth rates in the year 2010 with time trends since 1990 for selected countries: a systematic analysis and implications,” The Lancet, vol. 379, no. 9832, pp. 2162–2172, 2012, doi: 10.1016/s0140-6736(12)60820-4.

[7] M. M. Medvedev et al., “Development and validation of a simplified score to predict neonatal mortality risk among neonates weighing 2000 g or less (NMR-2000): an analysis using data from the UK and The Gambia,” (in eng), Lancet Child Adolesc Health, vol. 4, no. 4, pp. 299–311, Apr 2020, doi: 10.1016/s2352-4642(20)30021-3.

[8] N. J. Russell et al., “Patterns of antibiotic use, pathogens, and prediction of mortality in hospitalized neonates and young infants with sepsis: A global neonatal sepsis observational cohort study (NeoOBS),” (in eng), PLoS Med, vol. 20, no. 6, p. e1004179, Jun 2023, doi: 10.1371/journal.pmed.1004179.

[9] K. P. Mansoor et al., “Modified Sick Neonatal Score (MSNS): A Novel Neonatal Disease Severity Scoring System for Resource-Limited Settings,” (in eng), Crit Care Res Pract, vol. 2019, p. 9059073, 2019, doi: 10.1155/2019/9059073.

[10] B. Garg, D. Sharma, and N. Farahbakhsh, “Assessment of sickness severity of illness in neonates: review of various neonatal illness scoring systems,” (in eng), J Matern Fetal Neonatal Med, vol. 31, no. 10, pp. 1373–1380, May 2018, doi: 10.1080/14767058.2017.1315665.

[11] H. Bhandekar, S. Bansode Bangartale, and I. Arora, “Evaluating the Clinical Risk Index for Babies (CRIB) II Score for Mortality Prediction in Preterm Newborns: A Prospective Observational Study at a Tertiary Care Hospital,” (in eng), Cureus, vol. 16, no. 4, p. e58672, Apr 2024, doi: 10.7759/cureus.58672.

[12] D. K. Richardson, J. D. Corcoran, G. J. Escobar, and S. K. Lee, “SNAP-II and SNAPPE-II: Simplified newborn illness severity and mortality risk scores,” The Journal of Pediatrics, vol. 138, no. 1, pp. 92–100, 2001/01/01/ 2001, doi: 10.1067/mpd.2001.109608.

[13] S. Mahtab et al., “Causes of death identified in neonates enrolled through Child Health and Mortality Prevention Surveillance (CHAMPS), December 2016 –December 2021,” PLOS Global Public Health, vol. 3, no. 3, p. e0001612, 2023, doi: 10.1371/journal.pgph.0001612.

[14] L. Tibaijuka et al., “Incidence and predictors of preterm neonatal mortality at Mbarara Regional Referral Hospital in South Western Uganda,” (in eng), PLoS One, vol. 16, no. 11, p. e0259310, 2021, doi: 10.1371/journal.pone.0259310.

[15] J. B. Griffin, A. H. Jobe, D. Rouse, E. M. McClure, R. L. Goldenberg, and B. D. Kamath-Rayne, “Evaluating WHO-Recommended Interventions for Preterm Birth: A Mathematical Model of the Potential Reduction of Preterm Mortality in Sub-Saharan Africa,” (in eng), Glob Health Sci Pract, vol. 7, no. 2, pp. 215–227, Jun 2019, doi: 10.9745/ghsp-d-18-00402.

[16] B. T. Mmbaga, R. T. Lie, R. Olomi, M. J. Mahande, G. Kvåle, and A. K. Daltveit, “Cause-specific neonatal mortality in a neonatal care unit in Northern Tanzania: a registry based cohort study,” (in eng), BMC Pediatr, vol. 12, p. 116, Aug 7 2012, doi: 10.1186/1471-2431-12-116.

[17] World Health Organization. “SDG Target 3.2: Newborn and child mortality.” https://www.who.int/data/gho/data/themes/topics/indicator-groups/indicator-group-details/GHO/sdg-target-3.2-newborn-and-child-mortality (accessed 20 January 2024, 2024).

[18] World Health Organization, “Standards for improving the quality of care for small and sick newborns in health facilities.,” 2020.

[19] World Health Organization, “WHO recommendations for care of the preterm or low birth weight infant,” 2022. [Online]. Available: https://iris.who.int/bitstream/handle/10665/363697/9789240058262-eng.pdf

[20] J. J. Ho, P. Subramaniam, D. J. Henderson-Smart, and P. G. Davis, “Continuous distending pressure for respiratory distress syndrome in preterm infants,” (in eng), Cochrane Database Syst Rev, no. 2, p. Cd002271, 2002, doi: 10.1002/14651858.Cd002271.

[21] S. Martin, T. Duke, and P. Davis, “Efficacy and safety of bubble CPAP in neonatal care in low and middle income countries: a systematic review,” (in eng), Arch Dis Child Fetal Neonatal Ed, vol. 99, no. 6, pp. F495–504, Nov 2014, doi: 10.1136/archdischild-2013-305519.

[22] J. Myhre et al., “Effect of Treatment of Premature Infants with Respiratory Distress Using Low-cost Bubble CPAP in a Rural African Hospital,” Journal of Tropical Pediatrics, vol. 62, no. 5, pp. 385–389, 2016, doi: 10.1093/tropej/fmw023.

[23] A. B. Mwatha, M. Mahande, R. Olomi, B. John, and R. Philemon, “Treatment outcomes of Pumani bubble-CPAP versus oxygen therapy among preterm babies presenting with respiratory distress at a tertiary hospital in Tanzania-Randomised trial,” (in eng), PLoS One, vol. 15, no. 6, p. e0235031, 2020, doi: 10.1371/journal.pone.0235031.

[24] J. Carns et al., “National scale of neonatal CPAP to district hospitals in Malawi improves survival for neonates weighing between 1.0 and 1.3 kg,” (in eng), Arch Dis Child, vol. 107, no. 6, pp. 553–557, Jun 2022, doi: 10.1136/archdischild-2021-322964.

[25] J. Carns et al., “Neonatal CPAP for Respiratory Distress Across Malawi and Mortality,” Pediatrics, vol. 144, no. 4, 2019, doi: 10.1542/peds.2019-0668.

[26] U. M. Diala, B. O. Toma, D. D. Shwe, A. O. D. Ofakunrin, O. O. Diala, and C. John, “An assessment of improved outcomes using low-cost bubble CPAP in very low birthweight neonates in a Nigerian tertiary hospital,” Tropical Doctor, vol. 52, no. 4, pp. 503–509, 2022, doi: 10.1177/00494755221107461.

[27] C. Aneji et al., “Implementing bubble continuous positive airway pressure in a lower middle-income country: a Nigerian experience,” (in eng), Pan Afr Med J, vol. 37, p. 10, 2020, doi: 10.11604/pamj.2020.37.10.24911.

[28] A. Thukral, M. J. Sankar, A. Chandrasekaran, R. Agarwal, and V. K. Paul, “Efficacy and safety of CPAP in low- and middle-income countries,” (in eng), J Perinatol, vol. 36 Suppl 1, no. Suppl 1, pp. S21–8, May 2016, doi: 10.1038/jp.2016.29.

[29] G. de Carvalho Nunes et al., “Early Bubble CPAP Protocol Implementation and Rates of Death or Severe BPD,” (in eng), Pediatrics, vol. 154, no. 1, Jul 1 2024, doi: 10.1542/peds.2023-065373.

[30] P. Subramaniam, J. J. Ho, and P. G. Davis, “Prophylactic or very early initiation of continuous positive airway pressure (CPAP) for preterm infants,” (in eng), Cochrane Database Syst Rev, vol. 10, no. 10, p. Cd001243, Oct 18 2021, doi: 10.1002/14651858.CD001243.pub4.

[31] S. Dada et al., “Experiences with implementation of continuous positive airway pressure for neonates and infants in low-resource settings: A scoping review,” (in eng), PLoS One, vol. 16, no. 6, p. e0252718, 2021, doi: 10.1371/journal.pone.0252718.

[32] M. W. Kinshella et al., “Barriers and facilitators to implementing bubble CPAP to improve neonatal health in sub-Saharan Africa: a systematic review,” (in eng), Public Health Rev, vol. 41, p. 6, 2020, doi: 10.1186/s40985-020-00124-7.

[33] B. Ezenwa, P. Akintan, I. Fajolu, J. Ladele, and C. Ezeaka, “Bubble CPAP in the Management of Respiratory Distress Syndrome in Resource Constrained Settings: The Luth Experience,” Pediatric Oncall, vol. 13, no. 1, 2016, doi: 10.7199/ped.oncall.2016.11.

[34] K. E. Dickson et al., “Every Newborn: health-systems bottlenecks and strategies to accelerate scale-up in countries,” (in eng), Lancet, vol. 384, no. 9941, pp. 438–54, Aug 2 2014, doi: 10.1016/s0140-6736(14)60582-1.

[35] L. Tooke et al., “Limited resources restrict the provision of adequate neonatal respiratory care in the countries of Africa,” (in eng), Acta Paediatr, vol. 111, no. 2, pp. 275–283, Feb 2022, doi: 10.1111/apa.16050.

[36] E. von Elm, D. G. Altman, M. Egger, S. J. Pocock, P. C. Gøtzsche, and J. P. Vandenbroucke, “Strengthening the Reporting of Observational Studies in Epidemiology (STROBE) statement: guidelines for reporting observational studies,” (in eng), Bmj, vol. 335, no. 7624, pp. 806–8, Oct 20 2007, doi: 10.1136/bmj.39335.541782.AD.

[37] J. H. Cross et al., “Neonatal inpatient dataset for small and sick newborn care in low-and middle-income countries: systematic development and multi-country operationalisation with NEST360,” BMC Pediatrics, vol. 23, no. 2, p. 567, 2023/11/15 2023, doi: 10.1186/s12887-023-04341-2.

[38] P. Ayieko et al., “Characteristics of admissions and variations in the use of basic investigations, treatments and outcomes in Kenyan hospitals within a new Clinical Information Network,” (in eng), Arch Dis Child, vol. 101, no. 3, pp. 223–9, Mar 2016, doi: 10.1136/archdischild-2015-309269.

[39] R. E. Penzias et al., “Health facility assessment of small and sick newborn care in low-and middle-income countries: systematic tool development and operationalisation with NEST360 and UNICEF,” BMC Pediatrics, vol. 23, no. 2, p. 655, 2024/03/07 2024, doi: 10.1186/s12887-023-04495-z.

[40] R. E. Penzias et al., “Quantifying health facility service readiness for small and sick newborn care: comparing standards-based and WHO level-2 + scoring for 64 hospitals implementing with NEST360 in Kenya, Malawi, Nigeria, and Tanzania,” BMC Pediatrics, vol. 23, no. 2, p. 656, 2024/03/12 2024, doi: 10.1186/s12887-024-04578-5.

[41] R. E. Penzias et al., “Neonatal unit human resources: coverage for six cadres and trends for staff-to-baby ratios in 65 neonatal units implementing with NEST360 in Kenya, Malawi, Nigeria, and Tanzania,” Human Resources for Health, vol. 23, no. 1, 2025, doi: 10.1186/s12960-025-01031-1.

[42] UNICEF. NEST360. “Implementation Toolkit for Small and Sick Newborn Care.” https://www.newborntoolkit.org/ (accessed.

[43] World Health Organization, “Monitoring the Building Blocks of Health Systems: a Handbook of Indicators and their Measurement Strategies,” 2010, vol. 110. [Online]. Available: https://iris.who.int/handle/10665/258734

[44] E. Molyneux, Werdenberg, J., Liaghati-Mobarhan, S., Gicheha, E., & Langton, J, “Clinical Education Module: Bubble CPAP,” 07/2024 2024. [Online]. Available: https://nest360.org/wp-content/uploads/2024/07/Clinical-Module_bCPAP_FINAL-web_07-05-2024.pdf

[45] J. E. Lawn et al., “Small babies, big risks: global estimates of prevalence and mortality for vulnerable newborns to accelerate change and improve counting,” The Lancet, vol. 401, no. 10389, pp. 1707–1719, 2023/05/20/ 2023, doi: 10.1016/S0140-6736(23)00522-6.

[46] World Health Organization, “HIV strategic information for impact. Cascade data use manual: To identify gaps in HIV and health services for programme improvement.,” 2018. [Online]. Available: http://www.who.int/hiv/pub/toolkits/hiv-cascade-data-use-manual/en

[47] A. E. Greenberg, S. L. Hader, H. Masur, A. T. Young, J. Skillicorn, and C. W. Dieffenbach, “Fighting HIV/AIDS In Washington, D.C,” Health Affairs, vol. 28, no. 6, pp. 1677–1687, 2009, doi: 10.1377/hlthaff.28.6.1677.

[48] T. Tanahashi, “Health service coverage and its evaluation,” (in eng), Bull World Health Organ, vol. 56, no. 2, pp. 295–303, 1978.

[49] M. L. McNairy et al., “Use of a Comprehensive HIV Care Cascade for Evaluating HIV Program Performance: Findings From 4 Sub-Saharan African Countries,” (in eng), J Acquir Immune Defic Syndr, vol. 70, no. 2, pp. e44–51, Oct 1 2015, doi: 10.1097/qai.0000000000000745.

[50] N. Bamat, E. A. Jensen, and H. Kirpalani, “Duration of continuous positive airway pressure in premature infants,” (in eng), Semin Fetal Neonatal Med, vol. 21, no. 3, pp. 189–95, Jun 2016, doi: 10.1016/j.siny.2016.02.005.

[51] R. R. Mamidi and C. T. McEvoy, “Extending CPAP in stable preterm infants to increase lung growth and development as measured by pulmonary function testing,” Seminars in Perinatology, vol. 49, no. 5, p. 152059, 2025/08/01/ 2025, doi: 10.1016/j.semperi.2025.152059.

[52] B. H. Jena and M. M. Jaldo, “Determinants of neonatal mortality in sub-Saharan Africa: systematic review and meta-analysis of adverse newborn conditions,” (in eng), BMC Public Health, vol. 25, no. 1, p. 3058, Sep 24 2025, doi: 10.1186/s12889-025-24419-z.

[53] World Health Organization, “Thermal Control of the Newborn: a practical guide,” 1993. [Online]. Available: https://iris.who.int/server/api/core/bitstreams/63dd9e93-ea0a-4c5d-9213-0d5c5f4611a7/content.

[54] E. O. Ohuma et al., “National, regional, and global estimates of preterm birth in 2020, with trends from 2010: a systematic analysis,” (in eng), Lancet, vol. 402, no. 10409, pp. 1261–1271, Oct 7 2023, doi: 10.1016/s0140-6736(23)00878-4.

[55] J. Myhre et al., “Effect of Treatment of Premature Infants with Respiratory Distress Using Low-cost Bubble CPAP in a Rural African Hospital,” (in eng), J Trop Pediatr, vol. 62, no. 5, pp. 385–9, Oct 2016, doi: 10.1093/tropej/fmw023.

[56] J. Carns et al., “Impact of hypothermia on implementation of CPAP for neonatal respiratory distress syndrome in a low-resource setting,” (in eng), PLoS One, vol. 13, no. 3, p. e0194144, 2018, doi: 10.1371/journal.pone.0194144.

[57] J. Wainaina et al., “Hypothermia amongst neonatal admissions in Kenya: a retrospective cohort study assessing prevalence, trends, associated factors, and its relationship with all-cause neonatal mortality,” (in eng), Front Pediatr, vol. 12, p. 1272104, 2024, doi: 10.3389/fped.2024.1272104.

[58] F. H. A. Osier, J. A. Berkley, A. Ross, F. Sanderson, S. Mohammed, and C. R. J. C. Newton, “Abnormal blood glucose concentrations on admission to a rural Kenyan district hospital: prevalence and outcome,” Archives of Disease in Childhood, vol. 88, no. 7, pp. 621–625, 2003, doi: 10.1136/adc.88.7.621.

[59] E. McGoldrick, F. Stewart, R. Parker, and S. R. Dalziel, “Antenatal corticosteroids for accelerating fetal lung maturation for women at risk of preterm birth,” Cochrane Database of Systematic Reviews, no. 12, 2020, doi: 10.1002/14651858.CD004454.pub4.

[60] B. Turigye et al., “Beyond facility-based births: Is Uganda delivering effective maternal and newborn care? An analysis of the 2022 demographic health survey and 2023 harmonized health facility assessment survey,” PLOS Global Public Health, vol. 5, no. 10, p. e0004949, 2025, doi: 10.1371/journal.pgph.0004949.

[61] V. V. Ramaswamy et al., “ELBW and ELGAN outcomes in developing nations-Systematic review and meta-analysis,” (in eng), PLoS One, vol. 16, no. 8, p. e0255352, 2021, doi: 10.1371/journal.pone.0255352.

[62] S. G. Hundalani, R. Richards-Kortum, M. Oden, K. Kawaza, A. Gest, and E. Molyneux, “Development and validation of a simple algorithm for initiation of CPAP in neonates with respiratory distress in Malawi,” (in eng), Arch Dis Child Fetal Neonatal Ed, vol. 100, no. 4, pp. F332–6, Jul 2015, doi: 10.1136/archdischild-2014-308082.

[63] S. Oza, J. E. Lawn, D. R. Hogan, C. Mathers, and S. N. Cousens, “Neonatal cause-of-death estimates for the early and late neonatal periods for 194 countries: 2000-2013,” (in eng), Bull World Health Organ, vol. 93, no. 1, pp. 19–28, Jan 1 2015, doi: 10.2471/blt.14.139790.

[64] V. V. Ramaswamy, R. Devi, and G. Kumar, “Non-invasive ventilation in neonates: a review of current literature,” (in English), Frontiers in Pediatrics, Review vol. Volume 11 - 2023, 2023–November–28 2023, doi: 10.3389/fped.2023.1248836.

[65] K. Burgoine et al., “Impact of early continuous positive airway pressure in the delivery room (DR-CPAP) on neonates < 1500 g in a low-resource setting: a protocol for a pilot feasibility and acceptability randomized controlled trial,” Pilot and Feasibility Studies, vol. 10, no. 1, p. 126, 2024/10/04 2024, doi: 10.1186/s40814-024-01552-x.

[66] A. Napyo et al., “Acceptability of immediate CPAP for preterm infants in the delivery room to mothers, caregivers and healthcare workers in a low-resource setting: a qualitative study,” BMC Pediatrics, vol. 25, no. 1, 2025, doi: 10.1186/s12887-025-06055-z.

[67] Y. C. Cho et al., “Barriers and enablers to kangaroo mother care prior to stability from perspectives of Gambian health workers: A qualitative study,” Frontiers in Pediatrics, vol. 10, 2022, doi: 10.3389/fped.2022.966904.

[68] M. M. Medvedev et al., “Process and costs for readiness to safely implement immediate kangaroo mother care: a mixed methods evaluation from the OMWaNA trial at five hospitals in Uganda,” BMC Health Services Research, vol. 23, no. 1, p. 613, 2023/06/10 2023, doi: 10.1186/s12913-023-09624-z.

[69] W. I. K. S. Group, “Immediate “Kangaroo Mother Care” and Survival of Infants with Low Birth Weight,” New England Journal of Medicine, vol. 384, no. 21, pp. 2028–2038, 2021, doi: doi:10.1056/NEJMoa2026486.

[70] World Health Organization, “WHO recommendations on antenatal corticosteroids for improving preterm birth outcomes.,” 2022.

[71] L. Gibbs, L. Tooke, and M. C. Harrison, “Short-term outcomes of inborn v. outborn very-low-birth-weight neonates (<1 500 g) in the neonatal nursery at Groote Schuur Hospital, Cape Town, South Africa,” (in eng), S Afr Med J, vol. 107, no. 10, pp. 900–903, Sep 22 2017, doi: 10.7196/SAMJ.2017.v107i10.12463.

[72] M. Mukhtar-Yola and Z. Iliyasu, “A review of neonatal morbidity and mortality in Aminu Kano Teaching Hospital, northern Nigeria,” (in eng), Trop Doct, vol. 37, no. 3, pp. 130–2, Jul 2007, doi: 10.1258/004947507781524683.

[73] O. K. Oyedele, “Disparities and barriers of health facility delivery following optimal and suboptimal pregnancy care in Nigeria: evidence of home births from cross-sectional surveys,” BMC Women’s Health, vol. 23, no. 1, p. 194, 2023/04/25 2023, doi: 10.1186/s12905-023-02364-6.

[74] G. A. Kumar et al., “Neonatal complications and referral practices at birth: insights from a population-based study in the Indian state of Bihar,” BMJ Open, vol. 15, no. 7, p. e098408, 2025, doi: 10.1136/bmjopen-2024-098408.

[75] M. Kiputa, N. Salim, P. P. Kunambi, and A. Massawe, “Referral challenges and outcomes of neonates received at Muhimbili National Hospital, Dar es Salaam, Tanzania,” PLOS ONE, vol. 17, no. 6, p. e0269479, 2022, doi: 10.1371/journal.pone.0269479.

[76] M. L. Plummer, E. Calvello Hynes, J. Fogarty, N. Toro Polanco, and T. Reynolds, “Optimizing people’s movement across the health system: a scoping review of referral systems within a primary health care approach,” (in eng), Prim Health Care Res Dev, vol. 26, p. e94, Nov 17 2025, doi: 10.1017/s1463423625100546.

[77] A. Conde-Agudelo and J. L. Díaz-Rossello, “Kangaroo mother care to reduce morbidity and mortality in low birthweight infants,” Cochrane Database of Systematic Reviews, vol. 2017, no. 2, 2016, doi: 10.1002/14651858.cd002771.pub4.

