## Supplemental Files for "Respiratory support with Continuous Positive Airway Pressure in preterm neonates: an analysis of coverage and quality of care in 66 neonatal units in Kenya, Malawi, Nigeria and Tanzania implementing with the NEST360 Alliance"

**Supplementary Material**

**Additional File 1: STROBE Checklist.** List of items which should be included in reports for observational studies.

| | Item No | Recommendation | Page No |
|---|---|---|---|
| **Title and abstract** | 1 | (*a*) Indicate the study's design with a commonly used term in the title or the abstract | 2 |
| | | (*b*) Provide in the abstract an informative and balanced summary of what was done and what was found | 2-3 |
| **Introduction** | | | |
| Background/rationale | 2 | Explain the scientific background and rationale for the investigation being reported | 3-6 |
| Objectives | 3 | State specific objectives, including any prespecified hypotheses | 6-7 |
| **Methods** | | | |
| Study design | 4 | Present key elements of study design early in the paper | 6-8 |
| Setting | 5 | Describe the setting, locations, and relevant dates, including periods of recruitment, exposure, follow-up, and data collection | 6-8 |
| Participants | 6 | (*a*) *Cohort study*—Give the eligibility criteria, and the sources and methods of selection of participants. Describe methods of follow-up<br>*Case-control study*—Give the eligibility criteria, and the sources and methods of case ascertainment and control selection. Give the rationale for the choice of cases and controls<br>*Cross-sectional study*—Give the eligibility criteria, and the sources and methods of selection of participants | 7-9 |
| | | (*b*) *Cohort study*—For matched studies, give matching criteria and number of exposed and unexposed<br>*Case-control study*—For matched studies, give matching criteria and the number of controls per case | |
| Variables | 7 | Clearly define all outcomes, exposures, predictors, potential confounders, and effect modifiers. Give diagnostic criteria, if applicable | 8-11, Additional File 7 |

| | | | |
|---|---|---|---|
| Data sources/ measurement | 8* | For each variable of interest, give sources of data and details of methods of assessment (measurement). Describe comparability of assessment methods if there is more than one group | 7-10, Additional File 7 |
| Bias | 9 | Describe any efforts to address potential sources of bias | 9-10 |
| Study size | 10 | Explain how the study size was arrived at | 6-7 |
| Quantitative variables | 11 | Explain how quantitative variables were handled in the analyses. If applicable, describe which groupings were chosen and why | 8-10, Additional File 7 |
| Statistical methods | 12 | (*a*) Describe all statistical methods, including those used to control for confounding | 8-10, Additional File 7 |
| | | (*b*) Describe any methods used to examine subgroups and interactions | 8-10, Additional File 7 |
| | | (*c*) Explain how missing data were addressed | 9-10, Additional File 3,4,7 |
| | | (*d*) *Cohort study*—If applicable, explain how loss to follow-up was addressed<br>*Case-control study*—If applicable, explain how matching of cases and controls was addressed<br>*Cross-sectional study*—If applicable, describe analytical methods taking account of sampling strategy | |
| | | (*e*) Describe any sensitivity analyses | 9-10, Additional File 3,4 |

**Results**

| | | | |
|---|---|---|---|
| Participants | 13* | (a) Report numbers of individuals at each stage of study—eg numbers potentially eligible, examined for eligibility, confirmed eligible, included in the study, completing follow-up, and analysed | 11-12 |
| | | (b) Give reasons for non-participation at each stage | Figure 1 |
| | | (c) Consider use of a flow diagram | Figure 1 |
| Descriptive data | 14* | (a) Give characteristics of study participants (eg demographic, clinical, social) and information on exposures and potential confounders | Table 2 |

| | | | |
|---|---|---|---|
| | | (b) Indicate number of participants with missing data for each variable of interest | Additional File 3 |
| | | (c) *Cohort study*—Summarise follow-up time (eg, average and total amount) | |
| Outcome data | 15* | *Cohort study*—Report numbers of outcome events or summary measures over time | |
| | | *Case-control study*—Report numbers in each exposure category, or summary measures of exposure | |
| | | *Cross-sectional study*—Report numbers of outcome events or summary measures | 14-15 |
| Main results | 16 | (*a*) Give unadjusted estimates and, if applicable, confounder-adjusted estimates and their precision (eg, 95% confidence interval). Make clear which confounders were adjusted for and why they were included | 11-15 |
| | | (*b*) Report category boundaries when continuous variables were categorized | n/a |
| | | (*c*) If relevant, consider translating estimates of relative risk into absolute risk for a meaningful time period | n/a |
| Other analyses | 17 | Report other analyses done—eg analyses of subgroups and interactions, and sensitivity analyses | Additional Files 3-7 |
| **Discussion** | | | |
| Key results | 18 | Summarise key results with reference to study objectives | 15-19 |
| Limitations | 19 | Discuss limitations of the study, taking into account sources of potential bias or imprecision. Discuss both direction and magnitude of any potential bias | 19 |
| Interpretation | 20 | Give a cautious overall interpretation of results considering objectives, limitations, multiplicity of analyses, results from similar studies, and other relevant evidence | 15-20 |
| Generalisability | 21 | Discuss the generalisability (external validity) of the study results | 19 |
| **Other information** | | | |
| Funding | 22 | Give the source of funding and the role of the funders for the present study and, if applicable, for the original study on which the present article is based | 21 |

#### Additional File 2: Local ethical approval for the complex evaluation of the implementation of a small and sick newborn care package with NEST360

| Country | Protocol Title | LEC Protocol ID |
|---|---|---|
| **Kenya** | Using a Health Facility Assessment to Assess Quality of New Born Care in Kenya | MSU/DRPI/MUERC/00810/19 |
| **Malawi** | Using a Health Facility Assessment to Assess Quality of Newborn Care in Malawi | NHSRC 2463 |
| **Nigeria** | Quality Improvement Study of the Implementation of a Package of Trainings and Technologies for the Delivery of Comprehensive Newborn Care in Nigeria: A Multi-Country Study | **LUTH:** ADM/DCST/HREC/APP/3487<br>**UCH:** UI/EC/20/0713<br>**NHREC:** NHREC/01/01/2007 |
| **Tanzania** | Implementation study to improve the quality of comprehensive newborn care through introduction of the package of Newborn Essential Solutions and Technologies (NEST) in Tanzania | **IHI:** IHI/IRB/01-2021<br>**MUHAS:** MUHAS-REC-12-2019-072<br>**NIMR:** 3405 |

**Abbreviations**: LEC; Local Ethics Committee, ID; Identity, MSU; Michigan State University, DRPI; Disability Right Promotion International, MUERC; Maseno University Ethics Review Committee, NHSRC; National Health Science Research Committee, LUTH; Lagos University Teaching Hospital, UCH; University College Hospital, NHREC; National Health Research Ethics Committee, IHI; Ifakara Health Institute, MUHAS; Muhimbili University of Health and Allied Science, NIMR; National Institute for Medical Research

#### Additional File 3: CPAP Coverage by Eligibility Group with Depiction of Missingness in CPAP Administration Data (January 2021-December 2024)

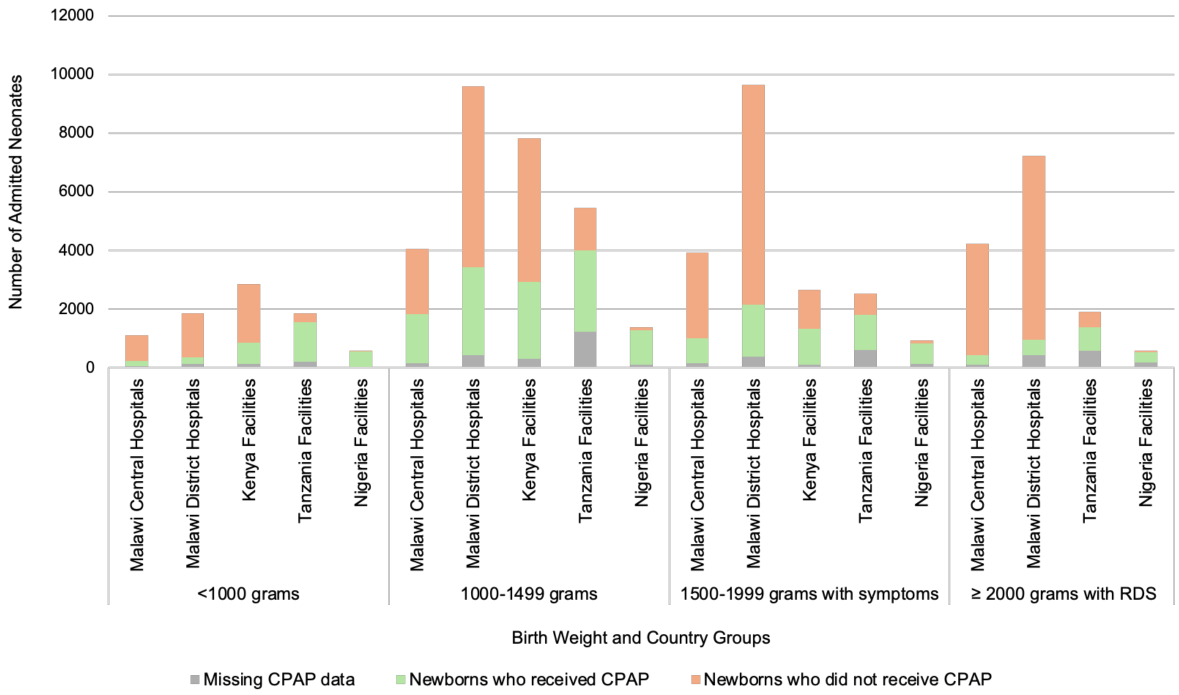


**Proportion of eligible neonates who received CPAP between January 2021 and December 2024.** Total N= 70,083 neonates.

The figure illustrates the breakdown of CPAP administration across eligibility groups and countries. The grey represents the number of neonates in each weight category with missing data for CPAP administration, the orange represents those who were eligible who did not receive CPAP, and the green represents neonates in each group who received CPAP. Of note, Kenya is not included for neonates >1999g due to differences in RDS diagnosis documentation.

**Additional File 4: Breakdown of CPAP administration data by facility (January 2021-December 2024)**

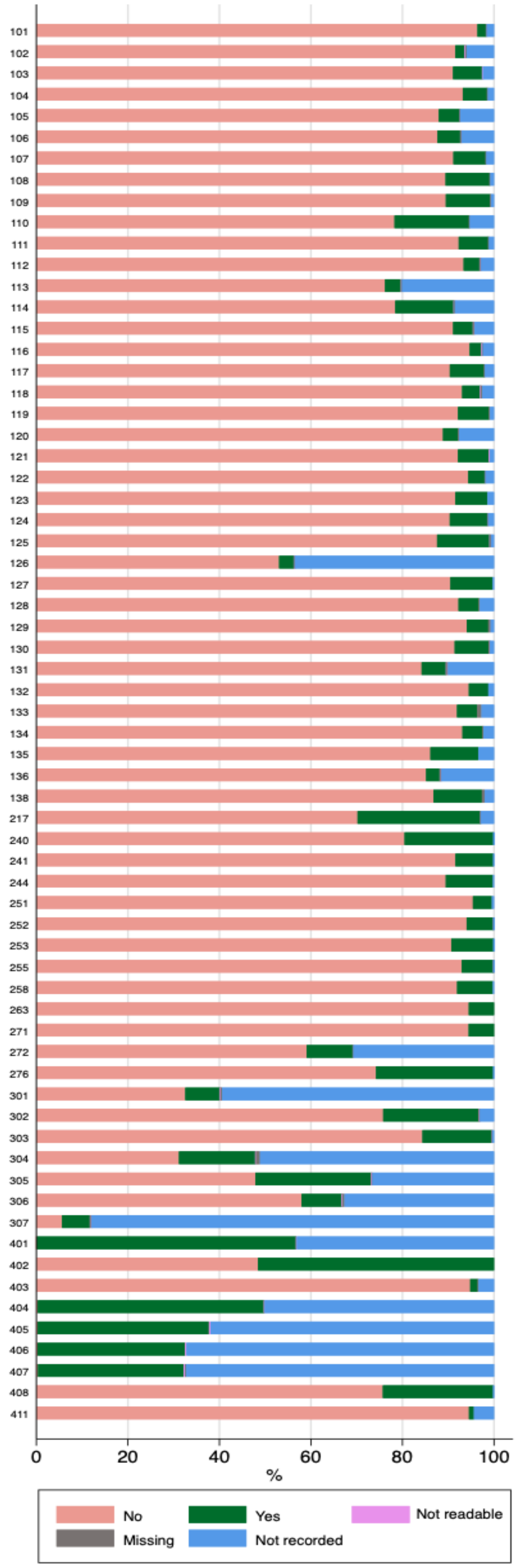


N= 375,255 neonatal admission records and 66 neonatal units. Classification of chart documentation for CPAP administration represented by colours in legend above.

**Additional File 5: Complete Case Analysis by Country and Weight Group (January 2021-December 2024)**

| Eligibility group | Country | Newborns who received CPAP | Total in group | Proportion with CPAP |
|---|---|---|---|---|
| Birth weight <1000 grams | Malawi Central Hospitals | 186 | 1,045 | 17.8 |
| | Malawi District Hospitals | 219 | 1,717 | 12.75 |
| | Kenya Facilities | 723 | 2,726 | 26.52 |
| | Tanzania Facilities | 1,360 | 1,644 | 82.73 |
| | Nigeria Facilities | 532 | 545 | 97.61 |
| Birth weight 1000-1499 grams | Malawi Central Hospitals | 1,685 | 3,907 | 43.13 |
| | Malawi District Hospitals | 3,000 | 9,181 | 32.68 |
| | Kenya Facilities | 2,642 | 7,526 | 35.1 |
| | Tanzania Facilities | 2,782 | 4,211 | 66.07 |
| | Nigeria Facilities | 1,170 | 1,280 | 91.41 |
| Birth weight 1500-1999 grams with symptoms | Malawi Central Hospitals | 836 | 3,782 | 22.1 |
| | Malawi District Hospitals | 1,773 | 9,256 | 19.16 |
| | Kenya Facilities | 1,219 | 2,558 | 47.65 |
| | Tanzania Facilities | 1,190 | 1,932 | 61.59 |
| | Nigeria Facilities | 716 | 816 | 87.75 |
| Birth weight ≥ 2000 grams with RDS | Malawi Central Hospitals | 323 | 4,128 | 7.82 |
| | Malawi District Hospitals | 524 | 6,797 | 7.71 |
| | Tanzania Facilities | 787 | 1,316 | 59.8 |
| | Nigeria Facilities | 339 | 394 | 86.04 |

#### Additional File 6: Additional Pooled Quality of Care Cascades

**A. Clinical quality cascade reflecting timing of CPAP initiation and duration of therapy for prophylactic CPAP, pooled across all countries (January 2021-December 2024).**

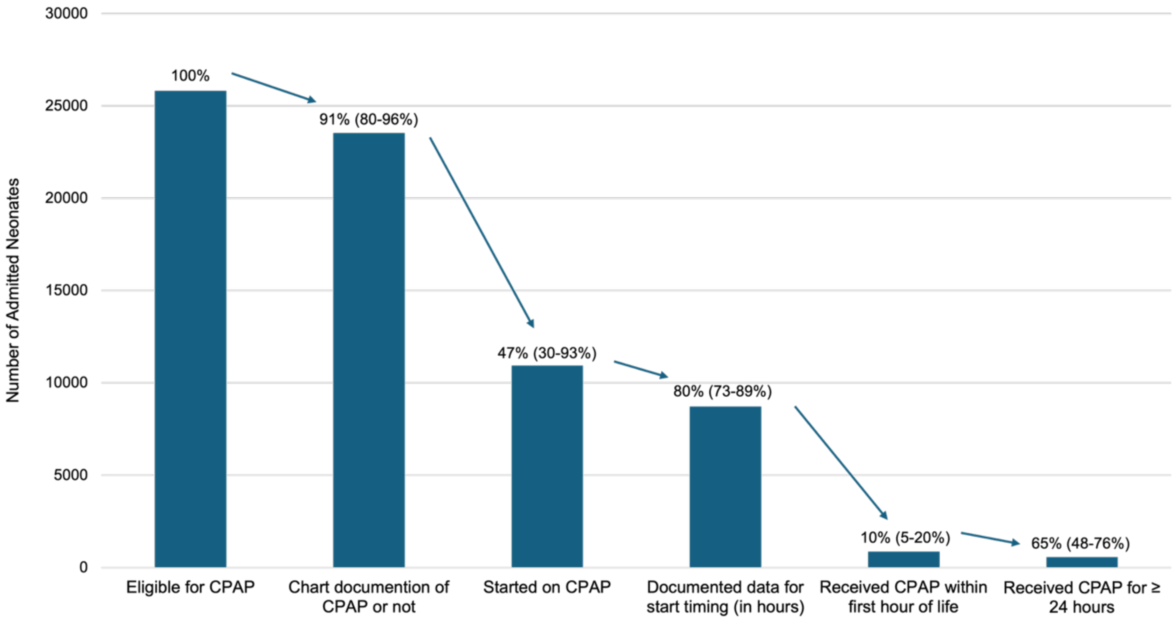


Initial n= 25,813 infants weighing <1500 grams. Results were pooled across all countries except Kenya, with the inter-country range illustrated in parentheses next to overall percentages above bars on the graph.

**B. Clinical quality cascade reflecting CPAP and IV fluid co-administration pooled across all countries (January 2021-December 2024).**

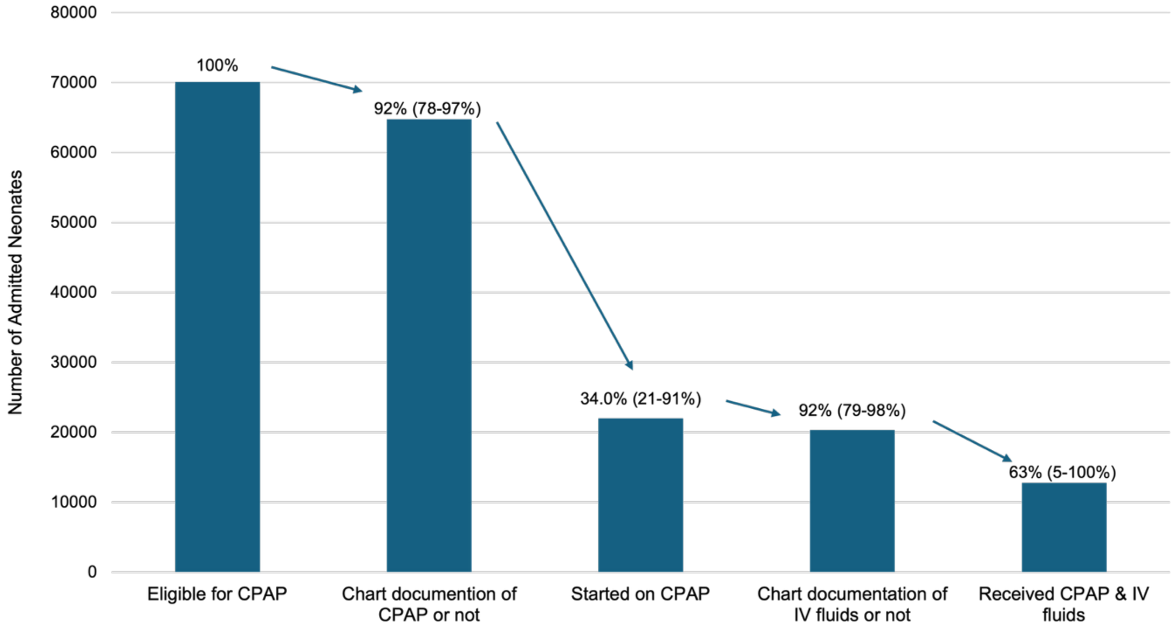


Initial n = 70,083. As in the other cascade, each subsequent indicator uses the preceding total as its denominator. Percentages represented above each bar, with inter-country ranges in parentheses.

**Additional File 7: Development of Severity Score and Outcome Model**

**Severity Score Conceptualization**

A clinical severity score for predicting survival among admitted neonates was needed to address mortality bias across multiple quality-of-care analyses. Many validated scores predict mortality or clinical illness (1-4) but were adaptable for this analysis. statistical analyses. The NMR-2000 is a validated score in LMIC setting adaptable for this analysis(2). We sought to create a parsimonious score using intrinsic variables at the We sought to create a parsimonious score using intrinsic variables at the individual level which do not include variables related to the intervention (or its indication) or hospital-level variables.

**Testing of Candidate Variables**

Missingness in proposed variables was explored during the analysis period (January 2021-December 2024). Variables were incorporated in different iterations in severity score models and tested for their statistical strength of association with survival. Candidate variables explored are detailed in the table below.

| NMR2000 variable used | Other variables considered | Variables Tested | Challenges |
|---|---|---|---|
| ***BWT/admission weight*** | | Gestational Age | Denominator circularity |
| ***Highest level of respiratory support in first 24 hours*** | | Not applicable | Numerator circularity |
| ***Pulse oximetry (SpO2) level on admission*** | | SpO2 on admission | |
| | ***Respiratory distress*** | Not applicable | Denominator circularity |
| | | Hypothermia on admission | |
| | ***Outborn*** | Outborn vs inborn | Binary, varies by facility |
| | ***Congenital anomalies*** | Discharge diagnosis of CA | Binary, varies by facility |
| | ***Age at admission*** | Age at admission | ~30% missing and co-linear with outborn |
| | ***Sex*** | Sex | No improvement in statistical association; should remain co-variate |

**Severity Categories**

Severity groupings are consistent with literature and/or WHO guidelines for each variable (5-9). Statistical tertiles based on input data not as informative, as the expected skew due to clinical characteristics among admitted babies helps predict actual severity. Statistical tertiles would remove this clinically meaningful skew and lead to regression to mean, as well as results which are sensitive but not specific.

| **Variable** | **Low Severity (1 point per each category)** | **Moderate Severity (2 points per each category)** | **High Severity (3 points per each category)** |
|---|---|---|---|
| **Gestational Age** | ≥32 weeks | ≥28 to <32 weeks | <28 weeks |
| **SpO2 on admission** | 90-100% | 80-89% | <80% |
| **Hypothermia on admission** | ≥36°C-37.5°C (mild hypothermia or normal temp) | 32.0°C–35.9°C | <32.0°C |

Lowest possible score = 3, highest possible score = 9

**Severity Scores Model Testing:**

Regression modelling performed to test scores' ability to predict survival in the NID dataset (January 2021-December 2024). Model 1 had slightly better overall likelihood fit for predicting survival. The AUC for Model 1 was 0.73 (95% CI 0.730-0.74) among all admissions and 0.74 (95% CI 0.734-0.74) for admissions <2500g.

These results were validated in the 2025 NID dataset, with an AUC of 0.734 across all admissions.

**Plot of Predicted Survival vs Severity Score (all admissions):**

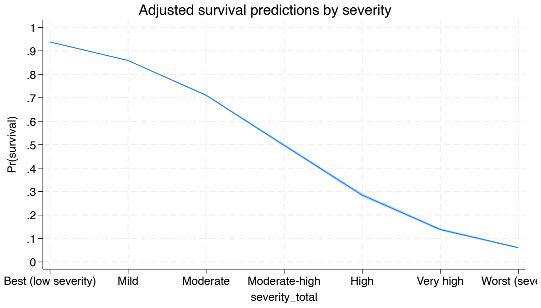


**Model 1**

N= 260,915, 95% CIs shaded around trend line

**Plot of Predicted Survival vs Severity Score (admissions<=2500g)**

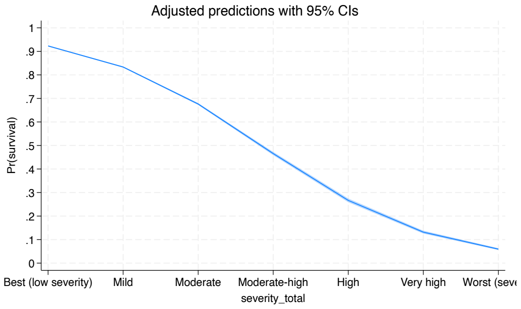


**Model 1**

N= 120,277, 95% CIs shaded around trend line

##### Severity Score Groupings

Scores are skewed due to clinical characteristics among admitted babies, and groups created to maintain clinically meaningful score categories:

**Low severity: 3-4 points**
Mostly low-risk features, maybe one moderate
**Moderate severity: 5-6 points**
Several moderate features or one high
**High severity: 7-9 points**
Multiple high-risk features (clinically concerning)

##### Original Regression Model

**Mixed-Effects Logistic Regression of Clinical Factors across A. all admitted, and B. all neonates receiving CPAP, (January 2021-December 2024).**

| Variable | A. Adjusted odds ratio (OR) and 95% CI among all admitted neonates (n=194,110; 66 neonatal units) | B. Adjusted odds ratio (OR) and 95% CI among neonates receiving CPAP (n=8,487; 53 neonatal units) |
|---|---|---|
| ***Temperature (ref: normal temperature)*** | | |
| Mild hypothermia (36°C–36.4°C) | 0.85** (0.81-0.89) | 0.90 (0.77-1.04) |
| Moderate or severe hypothermia (<35.9°C) | 0.45** (0.43-0.46) | 0.56** (0.50-0.63) |
| Elevated temperature or fever (≥37.5) | 0.61** (0.59-0.64) | 0.75* (0.61-0.92) |
| ***Birth location (ref: inborn)*** | | |
| Outborn | 0.41** (0.40-0.42) | 0.64** (0.57-0.72) |
| ***CPAP Administration (ref: none)*** | | |
| Received CPAP | 0.32** (0.31-0.33) | n/a |
| ***CPAP start time (ref ≤1 HOL)*** | | |
| >1 to 4 hours | n/a | 1.29* (1.07-1.56) |
| >4 to 12 hours | n/a | 0.90 (0.74-1.10) |
| >12 to 24 hours | n/a | 1.15 (0.94-1.42) |
| >24 hours | n/a | 1.56** (1.30-1.89) |
| ***CPAP duration (ref: <24 hours)*** | | |

| | | |
|---|---|---|
| 24–48 hours | n/a | 1.98** (1.75-2.25) |
| >48 hours | n/a | 3.22** (2.87-3.62) |
| ***Blood glucose (ref: Normoglycaemia 2.5–5.6 mmol/L)*** | | |
| Hypoglycaemia 1.65–2.5 mmol/L | 0.62** (0.55-0.69) | 0.64** (0.51-0.79) |
| Hypoglycaemia <1.65 mmol/L | 0.27** (0.23-0.31) | 0.25** (0.19-0.33) |
| Elevated glucose > 5.6 mmol/L | 0.95 (0.89-1.0) | 0.80* (0.66-0.95) |
| ***Signs of respiratory distress recorded (ref: none)*** | | |
| Respiratory distress | 0.35** (0.34-0.36) | 0.63** (0.56-0.71) |
| ***Maternal antenatal corticosteroids (ref: none)*** | | |
| Maternal corticosteroids given | 1.09* (1.04-1.15) | 1.03 (0.90-1.17) |
| ***Primary diagnosis (ref: prematurity)*** | | |
| Congenital Anomalies | 0.82** (0.77-0.88) | 0.82 (0.65-1.05) |
| Infection | 2.25** (2.1-2.4) | 1.08 (0.89-1.30) |
| Intrapartum-related | 1.05* (1.01-1.09) | 0.97 (0.80-1.18) |
| Jaundice | 6.21** (5.40-7.13) | 4.05** (2.05-8.01) |
| Other | 2.0** (1.91-2.09) | 1.01 (0.79-1.28) |

*A. Area under the curve (AUC)=0.79 (95%CI 0.786-0.791). LR test vs. logistic model: chibar2(01) = 2728, p-value <0.0001. Facility variance 0.19 (95% CI 0.13- 0.28).*
***p<0.001, *p<0.05, all others without statistical evidence of a relationship.*
*B. Area under the curve (AUC)=0.70 (95% CI 0.68-0.70). LR test vs. logistic model: chibar2(01) = 95, p-value <0.0001. Facility variance 0.19 (95% CI 0.11- 0.32).*
***p<0.001, *p<0.05, all others without statistical evidence of a relationship.*

**Additional File 8: Data Tools and Resources**

Health Facility Assessment (HFA) materials: https://nest360.org/project/hfa/

Neonatal Inpatient Dataset (NID) materials: https://nest360.org/project/nid/

**Additional File 9: Context Tracker Tool**

### NEST360 Neonatal Unit Context Tracker

#### WEEKLY REPORTING

| Question | Responses |
|---|---|
| **Electric power** | |
| Was there a loss of continuous electricity > 10 minutes on the neonatal unit in the last week?<br><br>*Indicate YES if there was a loss of electricity for more than 10 minutes that affected the neonatal unit in the last week. This can include a powercut from the grid or loss of backup power.* | Yes<br>No |
| Water Outage | |
| Was there a running water interruption lasting more than an hour on the neonatal unit in the last week? | Yes<br>No |
| **Human Resources** | |
| How many nurses are providing care in the neonatal unit?<br><br>*Do not include students in the count.*<br><br>**Staff providing care on the neonatal unit must be physically present at the hospital at the time to be counted.* | *Present today:____<br><br>Present last night:_____ |

#### MONTHLY REPORTING

| Question | Responses |
|---|---|
| **Human Resources**<br><br>**Staff providing care on the neonatal unit must be physically present at the hospital at the time to be counted.* | |
| How many doctors are providing care in the neonatal unit today? *Do not include students in the count.* | *Present today:<br><br>Present last night: |
| How many interns or residents are providing care on the neonatal unit today? *Do not include students in the count. This should only include fully qualified medical interns or residents.* | *Present today:<br><br>Present last night: |
| How many staff are currently on the neonatal unit roster? | Doctors:<br><br>Nurses: |

| Question | Responses | |
|---|---|---|
| Of those staff on the roster, how many have been formally trained on the national neonatal provider's course? | Doctors:<br><br>Nurses: | |
| How many biomedical technicians/engineers service the neonatal ward? *Do not include students in the count.* | *Present today:<br><br>Present last night: | |
| Of those biomedical technicians/engineers servicing the neonatal ward, how many have been formally trained on the neonatal devices? | | |
| Was there a nurse rotation in the last month. | Yes<br><br>No | |
| | If yes:<br><br>Date:<br><br>How many rotated out of the unit:<br><br>How many rotated into the unit: | |
| Was there a strike in the last month? | Yes<br><br>No | |
| | If yes<br><br>Cadre:<br><br>☐ Doctors<br>☐ Nurses<br>☐ Other (specify): ___________________<br>Start date:<br><br>End date: | |
| **Medical Devices** | | |
| **Device availability and functionality** | **<u>Total</u> devices in ward today**<br>*(Functional + not functional)* | **<u>Functional</u> devices in ward today**<br><br>*In depth functionality testing is not required. Functionality refers to the device, not consumable stockouts.* |
| Radiant warmer | | |
| Incubator | | |

| Question | Responses | | |
|---|---|---|---|
| Digital thermometer | | | |
| Suction pump | | | |
| Oxygen concentrator | | | |
| Oxygen tank | | | |
| Oxygen flow splitter | | | |
| CPAP | | | |
| Pulse oximeter | | | |
| Phototherapy | | | |
| Lightmeter | | | |
| Bilirubinometer | | | |
| Glucometer | | | |
| Hemoglobinometer | | | |
| Syringe pump | | | |
| Infusion pump | | | |
| Digital weighing scale | | | |
| Room heater | | | |
| **Consumables** | | | |
| **Consumables** | **Available today** | **Limited supply today** | **Not available today** |
| CPAP nasal cannula | | | |
| Oxygen nasal cannula (neonatal) | | | |
| Oxygen nasal cannula (infant) | | | |
| Suction pump catheters | | | |
| Glucometer strips | | | |
| Radiant warmer probes | | | |
| Incubator probes | | | |
| Pulse-oximeter probes | | | |
| BiliDx test strips | | | |
| **Medicines** | | | |

| Question | Responses | | |
|---|---|---|---|
| **Medicines** | **Available today** | **Limited supply today** | **Not available today** |
| Aminophylline | | | |
| Amoxicillin | | | |
| Ampicillin | | | |
| Caffeine | | | |
| Benzylpenicillin | | | |
| Gentamicin | | | |
| Vitamin K | | | |
| **Death Audits** | | | |
| How many neonatal deaths were audited in the last month? | Number: | | |
| How many audits had action plans developed? | Number: | | |
| **Neonatal Forms** | | | |
| Was there a stockout of neonatal forms in the last month? | Yes<br>No | | |
| | If yes:<br>Description: | | |
| **Disasters and Civil Unrest** | | | |
| Was there a natural disaster or civil unrest in the last month that affected the area covered by this neonatal unit? | Yes<br>No | | |
| | If yes:<br>☐ Flooding/heavy rains<br>☐ Heat wave<br>☐ Landslide<br>☐ Heat wave<br>☐ Cold wave<br>☐ Earthquake<br>☐ Drought<br>☐ Epidemic<br>☐ Civil unrest<br>☐ Other (specify):_____ | | |

| Question | Responses |
|---|---|
| | Start Date:<br><br>End Date (if ended): |
| | Description: |
| Was there any other event that impacted the ability to provide care on the neonatal unit? | Yes<br><br>No |
| *For example, this could include infrastructure challenges like broken windows, challenges with infection prevention supplies and devices.* | If yes:<br><br>Description |

#### References


1. Mansoor KP, Ravikiran SR, Kulkarni V, Baliga K, Rao S, Bhat KG, et al. Modified Sick Neonatal Score (MSNS): A Novel Neonatal Disease Severity Scoring System for Resource-Limited Settings. Crit Care Res Pract. 2019;2019:9059073.
2. Medvedev MM, Brotherton H, Gai A, Tann C, Gale C, Waiswa P, et al. Development and validation of a simplified score to predict neonatal mortality risk among neonates weighing 2000 g or less (NMR-2000): an analysis using data from the UK and The Gambia. Lancet Child Adolesc Health. 2020;4(4):299-311.
3. Russell NJ, Stöhr W, Plakkal N, Cook A, Berkley JA, Adhisivam B, et al. Patterns of antibiotic use, pathogens, and prediction of mortality in hospitalized neonates and young infants with sepsis: A global neonatal sepsis observational cohort study (NeoOBS). PLoS Med. 2023;20(6):e1004179.
4. Garg B, Sharma D, Farahbakhsh N. Assessment of sickness severity of illness in neonates: review of various neonatal illness scoring systems. J Matern Fetal Neonatal Med. 2018;31(10):1373-80.
5. World Health Organization. WHO recommendations for care of the preterm or low birth weight infant. 2022.
6. Jena BH, Jaldo MM. Determinants of neonatal mortality in sub-Saharan Africa: systematic review and meta-analysis of adverse newborn conditions. BMC Public Health. 2025;25(1):3058.
7. Nguyen TC, Madappa R, Siefkes HM, Lim MJ, Siddegowda KM, Lakshminrusimha S. Oxygen saturation targets in neonatal care: A narrative review. Early Hum Dev. 2024;199:106134.
8. Osier FHA, Berkley JA, Ross A, Sanderson F, Mohammed S, Newton CRJC. Abnormal blood glucose concentrations on admission to a rural Kenyan district hospital: prevalence and outcome. Archives of Disease in Childhood. 2003;88(7):621-5.
9. Ohuma EO, Moller AB, Bradley E, Chakwera S, Hussain-Alkhateeb L, Lewin A, et al. National, regional, and global estimates of preterm birth in 2020, with trends from 2010: a systematic analysis. Lancet. 2023;402(10409):1261-71.
